# Age-patterns of severity of clade I mpox in historically endemic countries

**DOI:** 10.1101/2024.04.23.24306209

**Authors:** Lilith K Whittles, Placide Mbala-Kingebeni, Neil M Ferguson

## Abstract

**Background:** The last two years have seen mpox emerge as a notable pandemic threat. The global 2022-23 outbreak of clade II mpox caused over 93,000 cases worldwide, while the 2023-24 epidemic of the more severe clade I virus in the Democratic Republic of Congo (DRC) has caused over 14,000 cases and over 650 deaths thus far. Of particular concern is that both case incidence and mortality in the DRC outbreak are concentrated in children.

However, quantification of age variation in severity for clade I infections has been lacking to date.

**Methods:** Using Bayesian binomial regression models, we analysed data from systematically reviewed clade I outbreaks to estimate case fatality ratios (CFR) by age, smallpox vaccination status and over time. We compared model predictive performance using leave-one-out cross-validation and compared our findings to the ongoing DRC outbreak.

**Findings:** The CFR had a near-reciprocal relationship with age, declining from 9.7% (95% credible interval: 6.9%-13.0%) among 5-year-olds to 1.2% (95%CrI: 0.3%-3.7%) by age 30. Accounting for vaccination status in addition to age did not improve model fit, but posterior parameter estimates suggest substantial vaccine-protection against death. Reanalysis incorporating cases from the ongoing DRC outbreak suggested less steep declines in severity with age and a protective effect of vaccination against death of 64% (95%CrI: - 3.4%-95.6%) and reduction in severity over time.

**Interpretation:** We provide estimates of the mpox clade I CFR in historically endemic settings by vaccine status, and find the highest risk of death in the youngest children.

**Funding:** Wellcome, NIHR, MRC, Community Jameel

## INTRODUCTION

Mpox is a viral zoonosis endemic to sylvatic rodent populations in West and Central Africa that has caused spill-over outbreaks among humans since its discovery in 1970 (1). Vaccination against smallpox, another orthopoxvirus, provides cross-protection against mpox infection (2). Smallpox vaccination campaigns ceased after its global eradication in 1980 and while vaccine-conferred protection is long-lasting, demographic turnover has reduced population-level vaccine-coverage worldwide, with marked regional variation (3). Susceptibility to orthopoxviruses is therefore concentrated in individuals born after 1980, who have never been infected with smallpox and are unlikely to have been vaccinated against it. With the waning of population immunity, mpox outbreaks have been increasingly frequent (1). In 2022 this culminated in a global outbreak comprising over 93,000 cases across 117 countries, during which sexual transmission was noted for the first time and was the key driver, with cases heavily concentrated among men who have sex with men (MSM) (4).

The mpox virus (MPXV) has been separated into distinct strains based on genetic differences: clade I (formerly known as the Congo Basin strain) and clade II (formerly the West African strain, a sub-clade of which, IIb, was responsible for the 2022 outbreak) (5). Severity of human MPXV infection differs by strain, the case fatality ratio (CFR) for clade I has been estimated at 10.6% (95%CI 8.4%-13.3%) compared with 4.6% (95%CI 2.1%-8.6%) for clade II, based on cases in endemic regions (1), while the CFR in the 2022-23 global clade IIb outbreak was much lower at <0.1% (6). Biological evidence supports strain-specific differences in virulence; in mice, clade I was estimated to be 1000x more severe than clade II based on fatal outcomes, with no deaths observed in sub-clade IIb (5).

In 2023 there were 14,626 suspected clade I mpox cases in the Democratic Republic of Congo (DRC), more than double the 6,216 cases recorded in 2020 (7,8). Sexual transmission of clade I mpox has been observed for the first time, with outbreaks among MSM networks and in commercial sex workers (7,8). However, unlike the clade IIb outbreak where over 95% of infections recording demographic data were among adult men (4), cases in the ongoing DRC epidemic have been concentrated in children, with 69% (3125/4538) of cases in the first 12 weeks of 2024 reported in under 15 year-olds, reflecting age-patterns observed in previous outbreaks in historically-affected countries (8). Furthermore, early evidence from DRC suggests that children under 15 years have accounted for 86% (254/296) of reported deaths in 2024, suggesting potential increased severity at young ages (8).

Age-specific variation in severity has important implications for understanding disease burden and how to best target interventions to reduce overall morbidity and mortality. However, there are currently no age-specific estimates of severity for mpox clade I. Furthermore, the extent to which age differences in severity can be explained by the concentration of vaccine-coverage at older ages is unclear.

Here, using data from systematically reviewed past outbreaks (1), we fitted mathematical models to quantify the relationship between the case fatality ratio (CFR) of mpox clade I and age at infection, the impact of vaccination status (either confirmed, or inferred based on birth date) on CFR, and looked for evidence of temporal changes in severity. We repeated the analysis incorporating information from the ongoing outbreak in DRC to assess the impact on our estimates.

## METHODS

### Data

We extracted data from a 2022 systematic review (1) on 550 case outcomes and year of symptom onset from studies reporting clade I outbreaks. Thirteen studies contained data on 125 individual cases including patient age and outcome (9–21). The individually reported cases ranged in age from eight months to 41 years. A further three studies reported 425 case outcomes grouped by age-band (22–24). The youngest age band in the grouped data was 0–3 years and the eldest was 40–69 years. Vaccination status of cases was recorded as ‘confirmed’ or ‘unvaccinated’ based on reported visual examination of vaccination scars. For the remaining cases we assigned vaccination status as ‘probable’ or ‘doubtful’ based on whether the patient was born before or after 1980, when routine smallpox vaccination ceased (3). We assessed the sensitivity of our findings to this assumption by repeating the analysis excluding all cases assigned as ‘probable’. We conducted supplementary analyses, assessing the impact of incorporating information from the ongoing outbreak in DRC, extracting 4538 age-grouped case outcomes (<1y, 1–5y, 5–14y, 15y+) reported in the DRC sitrep for the first 12 weeks of 2024 (11). We assigned ‘doubtful’ vaccination status to all cases from the ongoing DRC outbreak, Including the 1413 aged over 15, implicitly assuming that all cases were aged under 44 years (*i*.*e*. were born after 1980).

### Statistical analysis

In an exploratory analysis, we grouped the case data into five-year bands according to both age (*a*) *a*nd year of infection (*y*), as well as by vaccination status (*v*) *W*e compared patterns of cases (*C*_*j*_) *a*nd deaths (*D*_*j*_) *i*n each group *j =* {*a, y, v*}, and calculated crude estimates of the case fatality ratio by dividing the number of deaths by the number of cases CFR_*j*_ *= D*_*j*_/*C*_*j*_. We constructed exact 95% confidence intervals for the CFR using the by calculating the Clopper-Pearson interval.

We assessed the association between the CFR under each model, *p*_*M*_(*x*), and combinations of case covariates, including age (*x*_*a*_), year of onset (*x*_*y*_), and vaccination status *x*_*v*_ (where *x*_*v*_ *= 1* denotes probable or confirmed smallpox vaccination and *x*_*v*_ *= 0 d*enotes doubtful or no vaccination). When data was reported in age groups, we approximated the age of each case using the median age for each group, where reported, and otherwise calculated the midpoint of each age band. For the DRC data we assumed the average age of cases (and deaths) over the age of 15 was 34y, in line with national demography, and assessed the impact of our assumption in sensitivity analysis using a lower estimate of 21y.

We modelled the data using binomial regression and performed analyses both excluding and including the DRC sitrep data. We considered ten models (M1-M10) in total, allowing the CFR to depend on each of the three model covariates (age, vaccination status, year of onset) alone and in combination, and considering two different functional forms (logistic and Hill) for the dependency on age. For all models considered, the CFR can be represented in a generalised form:

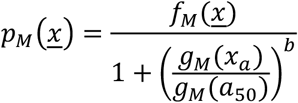

Here *b = 0 i*n the models where the CFR does not depend on age, 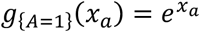 in the logistic models, and 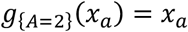 in the Hill function models. In models depending on age only, the numerator function *f*_{*V=0,Y=0*}_(*x*) *= θ*, where *θ i*s a parameter specifying the value of *p*_*M*_(*x*) at age zero in unvaccinated individuals. For models including vaccination status, *f*_{*V=1Y=0*}_(*x*) *= θ*(1 *− 𝜙x*_*v*_), where *𝜙 d*enotes the relative difference in CFR for vaccinated versus unvaccinated individuals. In models including year, 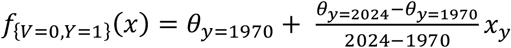, such that severity is assumed to vary linearly by year of onset between the two fitted endpoints. Last, in models including both year and vaccination status *f*_{*V*=1,*Y*=1}_ (*X*) = *f*_{*V*=1,*Y*=0}_( *f*_{*V*=0,*Y*=1}_ (*X*)).

The model parameters can be intuitively interpreted, such that *θ i*s the CFR at age zero for unvaccinated individuals, with year of onset *θ*_*y∈*{1970,2024}_ *w*here applicable; *𝜙 i*s the vaccine effectiveness against death given symptomatic infection; *a*_50_ *i*s the age by which the CFR has declined to half of its initial value of *θ*_*y*_; and *b c*ontrols the slope of the decline in CFR with age, where *b = 1 d*escribes a reciprocal relationship.

We fitted the models in a Bayesian framework using the R package brms v2.20.4 which serves as a frontend for the probabilistic programming language Stan (25,26). We used weakly informative uniform priors such that {*θ*_*y*_} *∼ U*(0,1), *a*_50_ *∼ U*(0,70), and *b ∼ U*(0,3). For the estimate of vaccine protection against death we adopted a deliberately conservative prior, *𝜙 ∼ U*(*−*1,1), allowing parameter values below zero to enable the model to detect whether the protective effect was statistically significant. We obtained posterior parameter estimates for each model using an adaptive Hamiltonian Monte Carlo no U-turn sampler comprising four MCMC chains of 5,000 iterations each. We assessed convergence of the chains by ensuring the Gelman-Rubin criterion was *≈*1 and the effective sample size (ESS) >1,000 for each parameter.

We compared the predictive performance of the ten fitted models via leave-one-out cross-validation, via the R package loo v2.6.0 (27) and applied a moment matching correction to the importance sampling for observations that were indicated as problematic, based on Pareto *k* diagnostic >0.7 (28).

The data and code needed to reproduce the analysis are freely available at: https://github.com/mrc-ide/mpox_clade_i_severity

## RESULTS

### Age and temporal trends in case data

Across the period 1970-2020, most data arose from the reports of outbreaks in DRC in the early 1980s and late 1990s (Figure 1A) (22,23). Cases over the whole period were concentrated in young age groups, with 38.5% (n=212/550) in children under 5 years old (Figure 1B). Most cases were unlikely to have been vaccinated against smallpox, with 55.5% (n=305) confirmed unvaccinated and 27.6% (n=152) unlikely to have been vaccinated due to being born after 1980. Only 9.3% (n=51) had confirmed smallpox vaccine scars. We assigned the remaining 7.6% (n=42) ‘probable’ vaccination status based on birth date prior to 1980. When cases were disaggregated into five-year age and time bands, there was a clear trend of cases occurring more frequently in adults in recent years, likely associated with the reductions in the proportion of adults protected by prior smallpox vaccination over time (Figure 1C).

**Figure 1.**
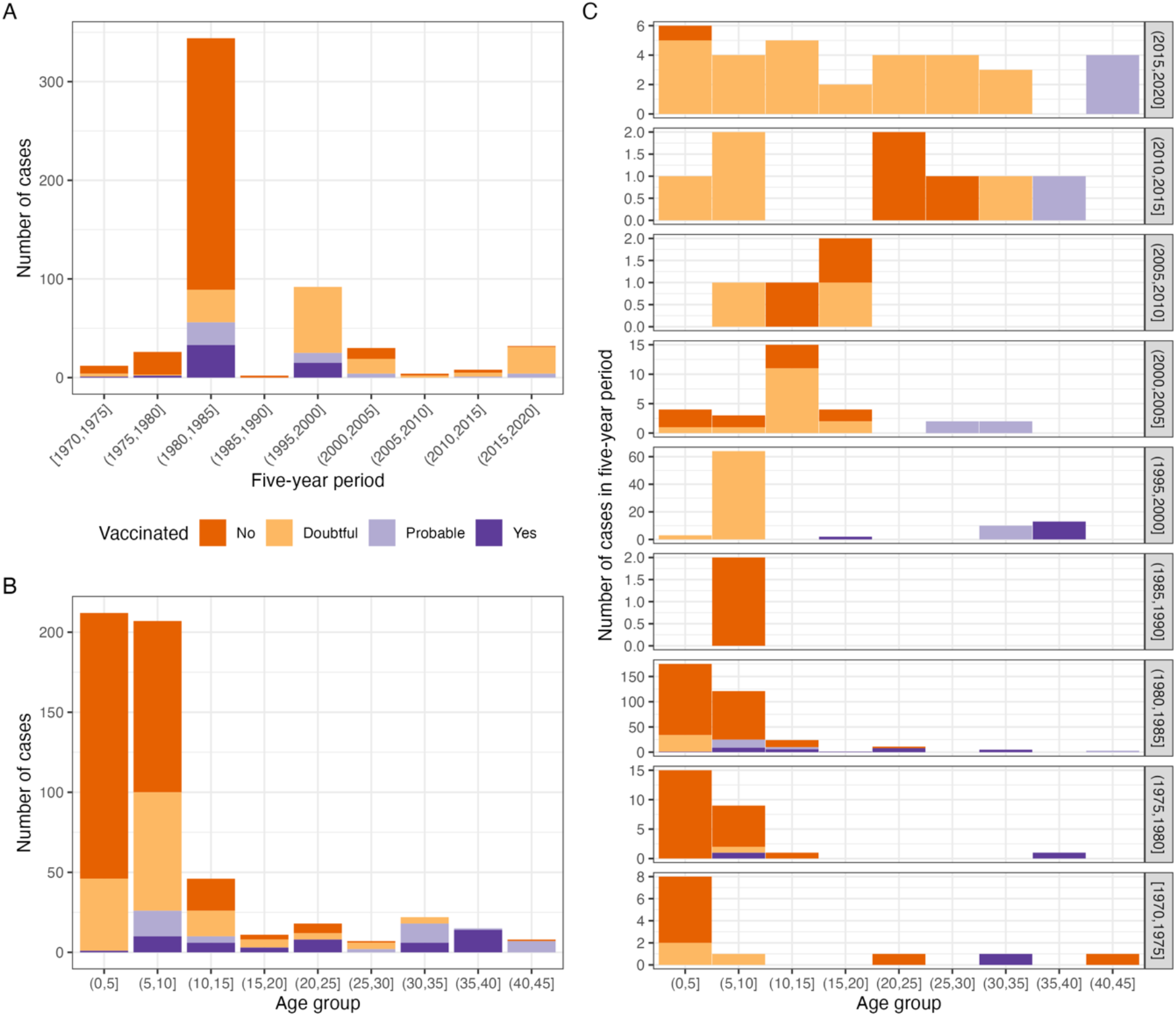
Number of cases recorded between 1970 and 2020 in studies identified by the Bunge et al. systematic review (1). A) in each five-year period over all ages, B) in each five-year age group over the whole period and C) split by both period and age. Stacked bars are coloured by vaccination status, where confirmed, and otherwise considered as ‘probable’ or ‘doubtful’ depending on whether an individual was born before or after 1980 (when routine smallpox vaccination ceased), respectively.

### Patterns of case severity by age and vaccination status

The overall crude CFR estimate from the pre-2021 case data was 9.5% (n=52/550; 95%CI 7.1%-12.2%). We performed preliminary analysis of age-trends in severity by stratifying the case data into broad age groups (children aged under five years, 5-14 years and “adults” aged 15 and over) which revealed more severe outcomes at younger ages. We estimated a crude CFR of 17.5% (n=37/212; 95%CI 12.6%-23.2%) in under 5 year-olds compared to 7.4% (n=13/175; 95%CI 4.0%-12.4%) in children aged 5-14 and 2.4% (n=2/85; 95%CI 0.3%-8.2%) in those aged 15 and over (Table 1)**Error! Reference source not found**.. A similar age-pattern of severity was also apparent in the 2024 DRC sitrep, although the decline in severity with age was less steep. The crude CFR in children under 5 was lower than the systematic review estimate with non-overlapping confidence intervals, at 10.5% (n=192/1832; 95%CI 9.1%-12.0%), declining to 4.8% (n=62/1293; 95%CI 3.7%-6.1%) in children aged 5-14 and 3.0% (n=42/1213; 95%CI 2.2%-4.0%) in those aged 15 and over.

**Table 1.**
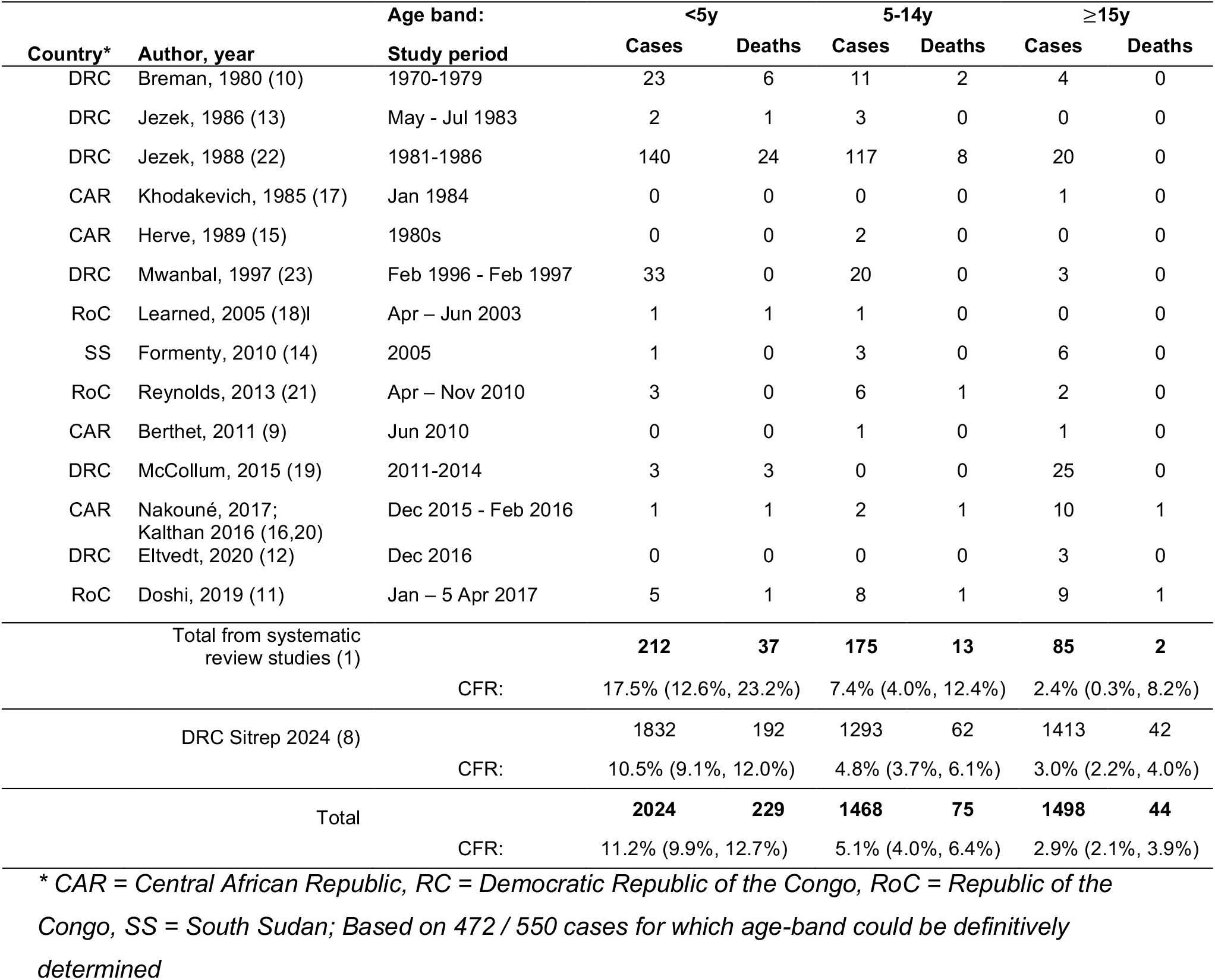
Analysis of crude CFR by age-group based on studies of clade I outbreaks included in Bunge et al. (1) only, and additionally incorporating 2024 DRC sitrep (8).

There were striking differences in case outcomes by vaccination status (Figure 2A); The crude CFR in the systematic review data was highest among unvaccinated individuals at 13.8% (n=42/263; 95%CI:10.1%-18.2%) (Figure 2B). By comparison, there were no recorded deaths among 51 cases in confirmed vaccinees. Indeed, the only fatal case that could potentially have been vaccinated was documented in a 40-year-old man, who was born three years before the cessation of smallpox vaccination in 1980 (11). The majority of cases in the systematic review data were recorded during the early 1980s outbreak in DRC (Figure 2C) (22), giving limited power to resolve temporal trends in CFR (Figure 2D).

**Figure 2.**
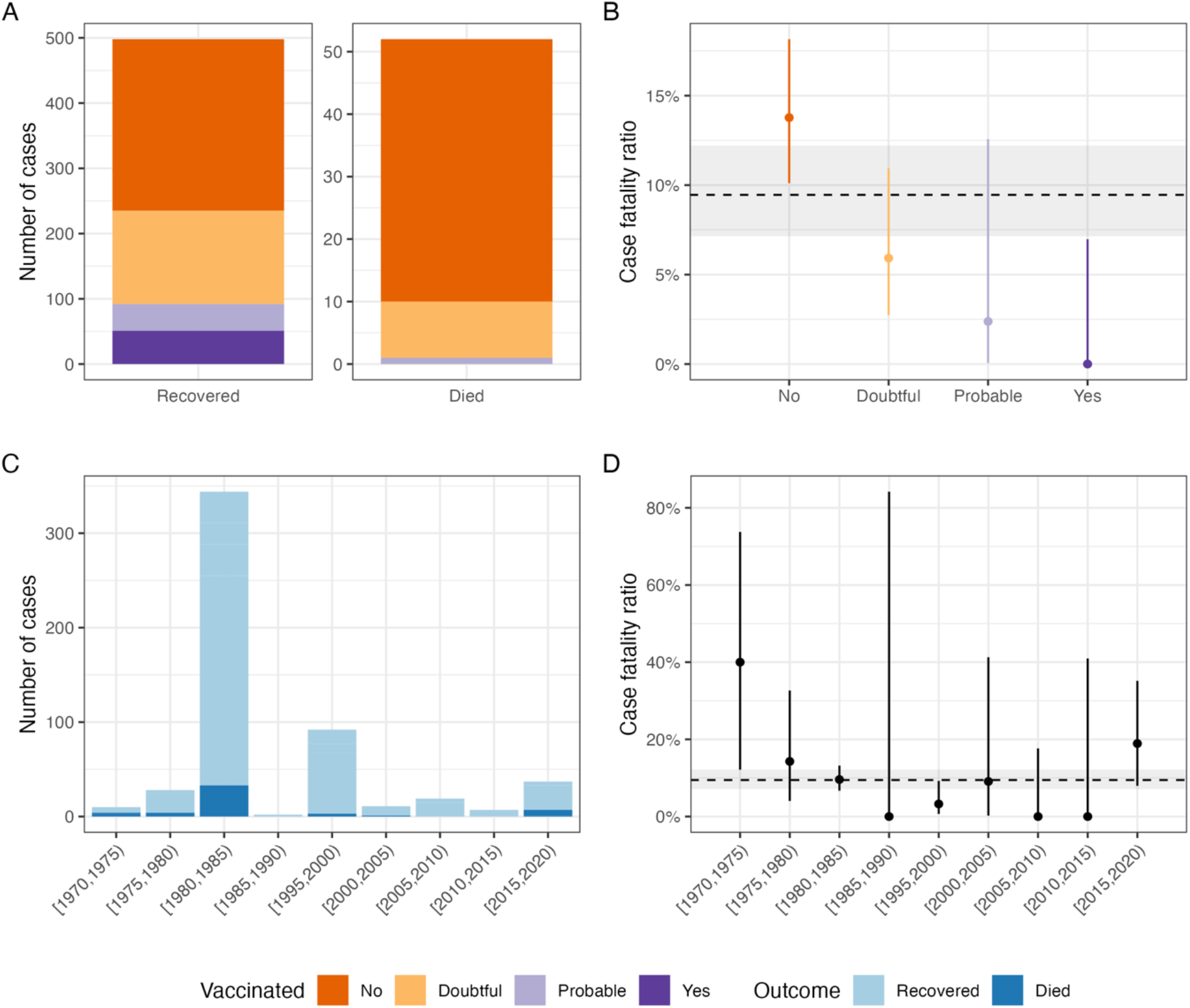
Case outcomes in studies identified by the Bunge et al. systematic review (1). A) by vaccination status with B) corresponding crude case fatality ratio (CFR); and C) in each five-year period since 1970 with D) corresponding crude CFR. In CFR plots (B,D), points depict the central estimate (deaths / cases) with error bars showing 95% exact binomial confidence intervals. Dashed horizontal lines and shaded bands show overall CFR (all ages, vaccine status and years) with 95% CI.

### Modelling the CFR

To formally characterise age variation in CFR while accounting for the potential confounding effects of temporal trends and differences in vaccine coverage by age, we fitted binomial regression models to the data reported in the systematic review (1). Leave-one-out cross validation confirmed that predictive performance was better for models assuming age dependence followed a Hill function (M7-10) than when logistic age dependence was assumed (M1,4-6) or models without age-dependence (M2-3), based on a difference in the expected log posterior density (ELPD) >4 (Table S1).

Increasing age was significantly associated with a decline in severity in all models. Modelling CFR as a Hill function of age only (M7), provided the best fit to data based on smallest ELPD, and capture. Comparing the modelled CFR by age-band to observed estimates from the data showed good correspondence (Figure S1). The posterior estimates of *a*_50_ *a*nd *b w*ere 2.9 (95% CrI 0.5 – 8.3) and 1.4 (95% CrI: 0.7 – 2.3) suggesting a near-reciprocal relationship between CFR and age. We estimate that CFR declines steeply from 26% (95%CrI 15% - 43%) at age 1 to 5% (95%CrI 3% - 8%) by age 10, falling to 1% (95%CrI 0% - 4%) by age 30 (Figure 3).

**Figure 3.**
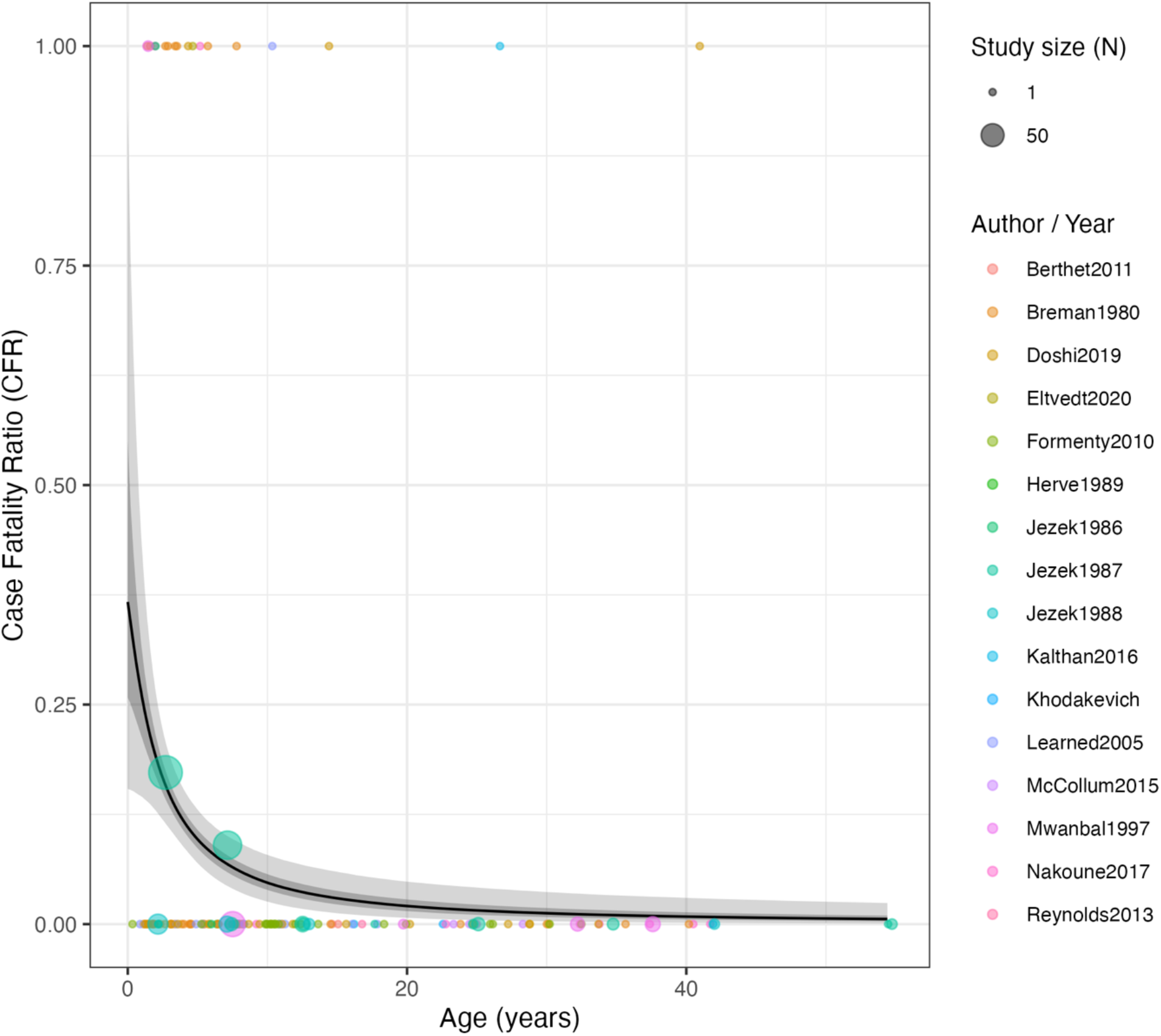
Estimated CFR by age for mpox clade I under Model 7 (Hill function, age-only). The solid line depicts the median posterior estimate, with shaded bands showing the 50% and 95% credible intervals. Points are coloured according to study and show the outcome of individual cases where reported (0 = recovery, 1 = death). Larger points depict the crude CFRs calculated in studies reporting outcome by age-group (plotted according to the middle age in the group range), with point size reflecting the number in each group.

Models that incorporated vaccination status in addition to age (M8,10) had similar ELPD (albeit with substantial uncertainty) to the simpler model including age only (M7), indicating that the inclusion of vaccine status did not substantially improve model fit. However, there was some suggestion of a protective effect of vaccination. Adding an effect of vaccination to the best fit model (M7) gave an estimated vaccine effectiveness (VE) against death of 44% (94%CrI: -55%, 94%), with 86% of posterior samples giving VE greater than zero (model M8 in Table S1). The posterior mean estimate was lowered by the deliberately conservative prior on vaccine effect, however the marginal posterior modal estimate was VE = 74% (Figure S 2). Repeating the analysis assuming a more biologically plausible *𝜙 ∼ U*[0,1] *p*rior gave an estimated vaccine effectiveness (VE) against death of 57% (94%CrI: 6%, 95%).

While models incorporating year of onset had similar ELPD values to the best-fitting age-only model (M7), there were insufficient data in recent years to be informative about changes in the CFR over time, with little evidence to distinguish the estimate of *θ*_*y*=2024_ from its prior (Figure S 2).

We assessed the sensitivity of our results to the assumption that individuals born before 1980 whose vaccine status was unknown were ‘probable’ vaccinees. Refitting the models excluding these 42 cases (including one death) produced similar ELPD estimates for all Hill function models, however the model incorporating vaccination (M8) had the lowest value overall, with similar parameter estimates and quality of fit to the main results (Table S2; Figure S3; Figure S4). While the analysis was underpowered to provide a statistically significant estimate of vaccine protection against death, the posterior was maximized at VE = 88% (Figure S 5).

### Analysis including data from DRC sitrep

We repeated model parameter estimation incorporating the data on mpox cases and deaths by age band (<1y, 1-4y, 5-14y, 15y+) reported in the 2024 DRC sitrep. Inclusion of the sitrep data changed the preferred model; Leave-one-out cross validation selected Model 10, where CFR was modelled as a Hill function of age and depended on both vaccination status and calendar time (Figure 4). The magnitude of the age-effect was diminished compared with the model fit based on historic data alone: the inferred CFR at age 0 was lower at 23% (95% CrI: 13% - 40%) in 1970 falling to 14% (95% CrI: 10% - 22%) in 2024, compared with 42% (95% CrI 15% - 93%) when the sitrep data was not included (Table S 3). Estimated vaccine efficacy against death increased to a mean of 64% (95% CrI: -2% - 96%) with 97% posterior support that vaccine efficacy was >0, with the posterior maximised at VE = 82% (Figure S 6). In addition, the estimate of *a*_50_ *i*ncreased to 6.8y (95% CrI: 1.6y – 12.9y) and that of *b*, the slope of the Hill function, decreased to 0.91 (95% CrI: 0.57– 1.28) (Table S 3)

**Figure 4.**
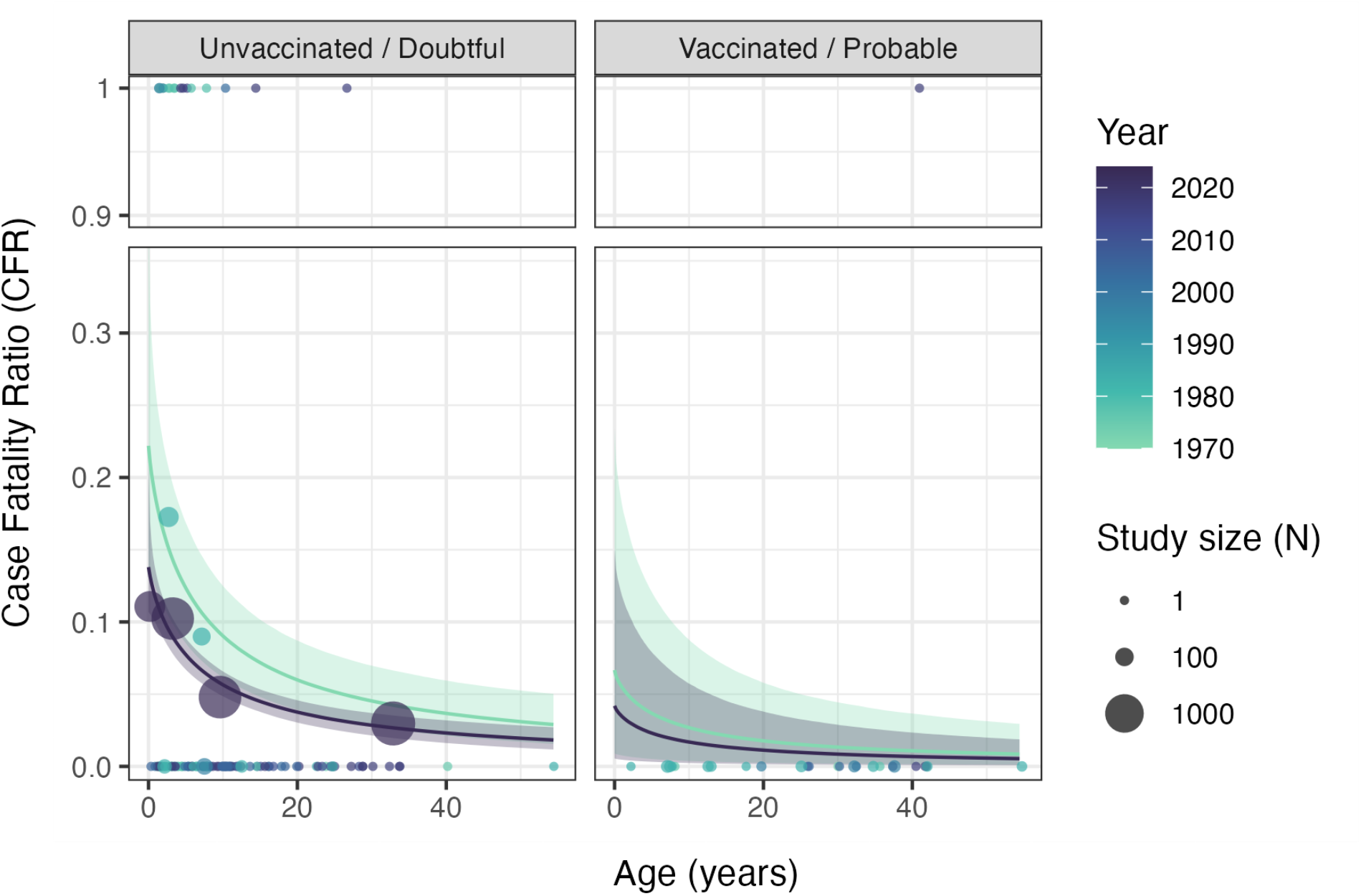
Estimated CFR by age, inferred vaccination status, and year of onset for mpox clade I under Model 10 (Hill, age, vaccination status, year) based on data incorporating the DRC 2024 sitrep. Solid lines depict the median posterior estimate for the posterior mean CFR at each age inferred for 1970 and 2024, with shaded bands showing the 50% and 95% credible intervals. Points are coloured according to year of outbreak and show the outcome of individual cases where reported (0 = recovery, 1 = death). Larger points depict the crude CFRs calculated in studies reporting outcome by age-group (plotted according to the median age in the group, where reported and mid-point of the age range otherwise), with point size reflecting the number in each group.

We assessed the sensitivity of these results to uncertainty around the age of adult cases in the 2024 DRC outbreak. In the analysis presented above, we assumed that the mean age of cases in the 15+ age group was 34y (in line with DRC demography). Reducing the average age of adult cases to 21y and repeating the analysis produced qualitatively unchanged results in terms of model selection and parameter estimates for the better fitting Hill function models (Table S4; Figure S8; Figure S 9; Figure S10).

Table 2 tabulates CFR estimates for two of the best fitting model variants from the main analysis and the best fitting model from the supplementary analysis which also included the 2024 DRC sitrep data.

**Table 2.**
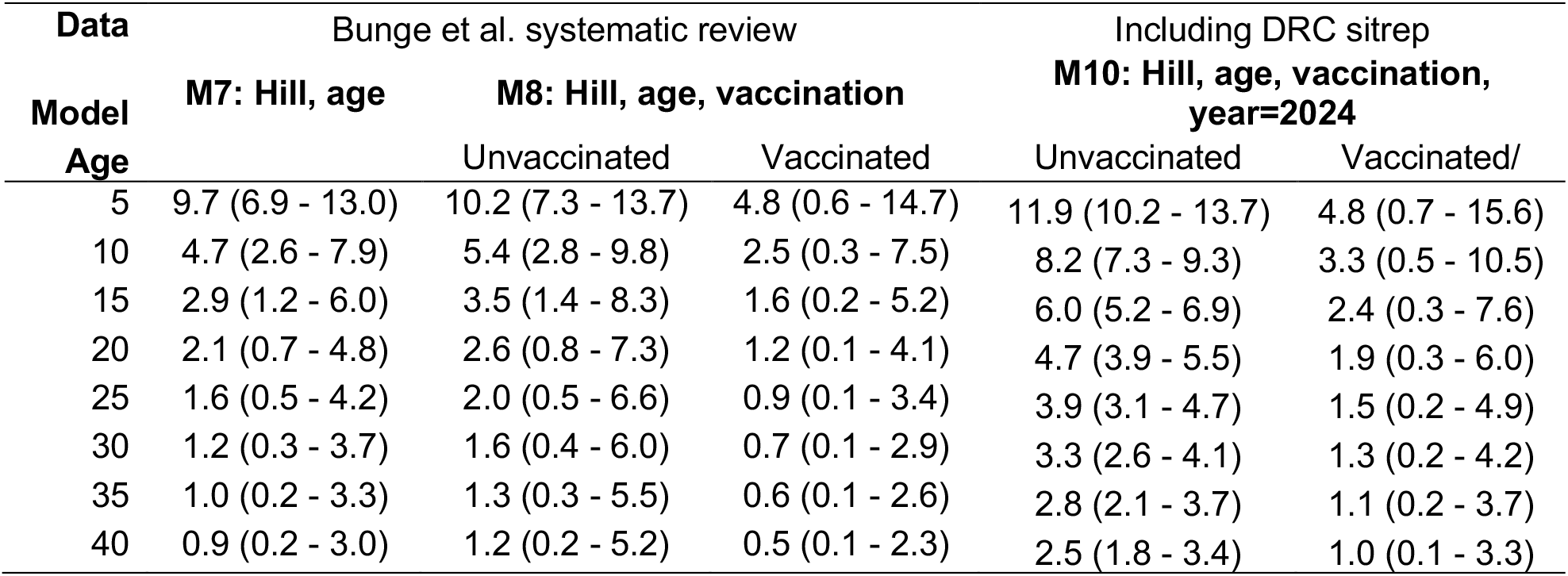
Modelled clade I case fatality ratio (CFR; %) by age, based on posterior mean and 95% credible interval for models M7 (Hill function, age-only) and M8 (Hill function, age and vaccination status) for the main analysis (including data from the Bunge et al. systematic review) and M10 for the analysis additionally incorporating data from the 2024 DRC sitrep, presenting CFR for 2024.

| Model<br>Age | Bunge et al. systematic review |  |  | Including DRC sitrep |  |
| --- | --- | --- | --- | --- | --- |
|  | M7: Hill, age | M8: Hill, age, vaccination |  | M10: Hill, age, vaccination, year=2024 |  |
|  |  | Unvaccinated | Vaccinated | Unvaccinated | Vaccinated/ |
| 5 | 9.7 (6.9 - 13.0) | 10.2 (7.3 - 13.7) | 4.8 (0.6 - 14.7) | 11.9 (10.2 - 13.7) | 4.8 (0.7 - 15.6) |
| 10 | 4.7 (2.6 - 7.9) | 5.4 (2.8 - 9.8) | 2.5 (0.3 - 7.5) | 8.2 (7.3 - 9.3) | 3.3 (0.5 - 10.5) |
| 15 | 2.9 (1.2 - 6.0) | 3.5 (1.4 - 8.3) | 1.6 (0.2 - 5.2) | 6.0 (5.2 - 6.9) | 2.4 (0.3 - 7.6) |
| 20 | 2.1 (0.7 - 4.8) | 2.6 (0.8 - 7.3) | 1.2 (0.1 - 4.1) | 4.7 (3.9 - 5.5) | 1.9 (0.3 - 6.0) |
| 25 | 1.6 (0.5 - 4.2) | 2.0 (0.5 - 6.6) | 0.9 (0.1 - 3.4) | 3.9 (3.1 - 4.7) | 1.5 (0.2 - 4.9) |
| 30 | 1.2 (0.3 - 3.7) | 1.6 (0.4 - 6.0) | 0.7 (0.1 - 2.9) | 3.3 (2.6 - 4.1) | 1.3 (0.2 - 4.2) |
| 35 | 1.0 (0.2 - 3.3) | 1.3 (0.3 - 5.5) | 0.6 (0.1 - 2.6) | 2.8 (2.1 - 3.7) | 1.1 (0.2 - 3.7) |
| 40 | 0.9 (0.2 - 3.0) | 1.2 (0.2 - 5.2) | 0.5 (0.1 - 2.3) | 2.5 (1.8 - 3.4) | 1.0 (0.1 - 3.3) |

## DISCUSSION

Our analysis provides strong evidence for variation in the CFR of mpox clade I by age in endemic regions. Modelling based on data included in the Bunge et al. systematic review suggests a near-reciprocal relationship between the CFR and age, with estimates of 9.7% (95% CrI 6.9% - 13.0%) for children aged 5, to 1.2% (95% CrI 0.3% - 3.7%) by age 30. While incorporating probable smallpox vaccination status in addition to age did not improve the model’s predictive ability, posterior estimates indicate protection against death for individuals who are vaccinated but nevertheless become symptomatically infected with mpox, although our analysis of the scant historical data was statistically underpowered with broad credible intervals overlapping zero.

Cases and deaths from the DRC epidemic reported in the first 12 weeks of 2024 imply a CFR of 10.5% (n=192/1832; 95% CI: 9.1%-12.0%) in <5s, 4.8% (n=62/1293; 95% CI: 3.7%- 6.1%) in 5-14 year olds, and 3.0% (n=42/1413; 95% CI: 2.2%- 4.0%) in over 15s (8). Reanalysis of the CFR combining these data with the earlier data collated by Bunge et al. showed age was still the most important determinant of severity. However, the magnitude of age differences was diminished compared with the analysis of data from the systematic review. With the expanded dataset, the model incorporating vaccination status and calendar time in addition to age was preferred over the model of age only. There was strong posterior support for a protective effect against death, with a vaccine efficacy estimate of 64% (95% CrI: -2% - 96%). In addition, there was support for a reduction in CFR between 1970 and 2024, although this was driven entirely by the 2024 outbreak data. The cause for the reduction in severity is unclear with potential contributing factors including differences in case ascertainment, improvements in population health and/or access to care, or changing virulence as the clade evolves, with further work needed to attribute causality and implications.

A key strength of our analysis include is its synthesis of evidence from systematically identified outbreaks within a robust Bayesian framework incorporating model selection to provide the first quantitative estimates of the CFR for clade I mpox in endemic settings. However, there are also important limitations. Transmission of mpox clade I is concentrated in central Africa and has not caused major outbreaks outside of this endemic region to date. As such, our analysis is restricted to cases from DRC, Republic of Congo, Central African Republic, and South Sudan. The level of surveillance, case ascertainment, prevalence of co-morbidities, and access to healthcare vary substantially between settings worldwide, so estimates of severity presented here are not necessarily transferrable to other contexts. We did not have data on underlying heterogeneities in susceptibility to severe mpox outcomes, which may be associated with age. Qualitatively, case reports of clade I infections reveal co-morbidities, such as measles co-infection, and preventable exacerbating conditions, such as dehydration and bacterial superinfection of lesions, suggesting wide scope for improvement of outcomes with appropriate care (16,20,22).

In the absence of genomic evidence, we considered cases to part of clade I based on location, in accordance with the systematic review underlying the analysis and geographical spread described by WHO (1). Transmission of clade II in clade I endemic areas could therefore lead to misspecification, although prior to the 2022 clade IIb outbreak this is likely to have been minimal.

The interpretation and applicability of our CFR estimates depends upon both case definition and ascertainment, which will vary depending on reporting system, e.g. via surveillance or active epidemiological investigation. Severe cases, especially those with fatal outcomes, are more likely to be identified than mild infections, which may lead to CFR being over-estimated. Furthermore, if reporting of severe outcomes is associated with age, for example if deaths in children are more likely to be reported than deaths in adults, the concentration of severity at young ages may be overestimated, however it is equally likely that case ascertainment overall is higher in children. The data underlying our analysis were limited and restricted to the details provided in published case reports, with the quantity and quality of data varying between outbreaks and locations. Case ascertainment in the current DRC epidemic is unclear with the majority of cases being suspected but unconfirmed; due to diagnostic capacity constraints, only around 10% of suspected cases are sampled for laboratory analysis. As such, some suspected mpox cases could be misattributed, leading to potential inclusion of other diseases that cause skin lesions, such as chickenpox, in the case count. Furthermore, as the DRC epidemic is ongoing, right-censoring of case-outcomes in the naïve estimator (29) may lead to CFR being underestimated – perhaps explaining the apparent reduction in CFR over time seen in our analysis which included 2024 DRC data in the case count.

Smallpox vaccination status was confirmed in only 65% of case reports in the systematic review and none of the 2024 DRC sitrep cases, so we inferred probable vaccination status based on birth dates. The implicit assumption that all birth cohorts before 1980 were vaccinated likely overestimates coverage in older age groups, as it exceeds the 80% target coverage set by WHO during its smallpox vaccination campaign (3), we addressed this limitation by including a sensitivity analysis excluding the 42 individuals born before 1980 whose vaccine status was unconfirmed, which favoured the model including age and vaccination status (M8) over age alone (M7), but otherwise made little qualitative difference to our results. Furthermore, misclassifying fatal case outcomes as being vaccinated lowers estimates of vaccine efficacy, making the assumption conservative in this respect.

While data from the ongoing DRC outbreak suggest a reduction in the CFR over time, an increase in sexual transmission of clade I mpox may adversely impact future trends in severity. During the 2022 clade IIb outbreak, people living with HIV were more likely to be infected with mpox, and those with advanced HIV infection were more likely to suffer severe mpox disease and death, indicating increased severity due to immune compromise (30). HIV prevalence is <1% in DRC (among 83% of people who know their status) but this rises to 7% in key populations (7), and we currently lack clade I CFR estimates in immunocompromised patients.

In conclusion, age dependent severity, together with the concentration of susceptibility to mpox infection at young ages due to lack of prior infection and the discontinuation of smallpox vaccination, raises important questions for future epidemic response. Despite mounting reports of sexual transmission of clade I infection in the ongoing DRC outbreak, the majority of cases are still reported in young children, suggestive of a generalised epidemic with sustained community transmission. Additional epidemiological studies backed by mathematical modelling are urgently needed to help decision-makers understand the interaction between drivers of transmission and the age-patterns of severity we present here, and their implications for targeting future vaccination campaigns.

## Data Availability

All data and code to reproduce the analysis are available at https://github.com/mrc-ide/mpox_clade_i_severity

## Contributors

The study was conceived by LKW. Study data was extracted by LKW. The analysis was designed and implemented by LKW with input from NMF. The report was drafted by LKW and revised by all authors. All authors had access to the data and accept responsibility for submission for publication.

## License

For the purpose of open access, the author has applied a ‘Creative Commons Attribution (CC BY) licence to any Author Accepted Manuscript version arising.

## Supplementary Appendix

### Main analysis: systematic review data

**Figure S1.**
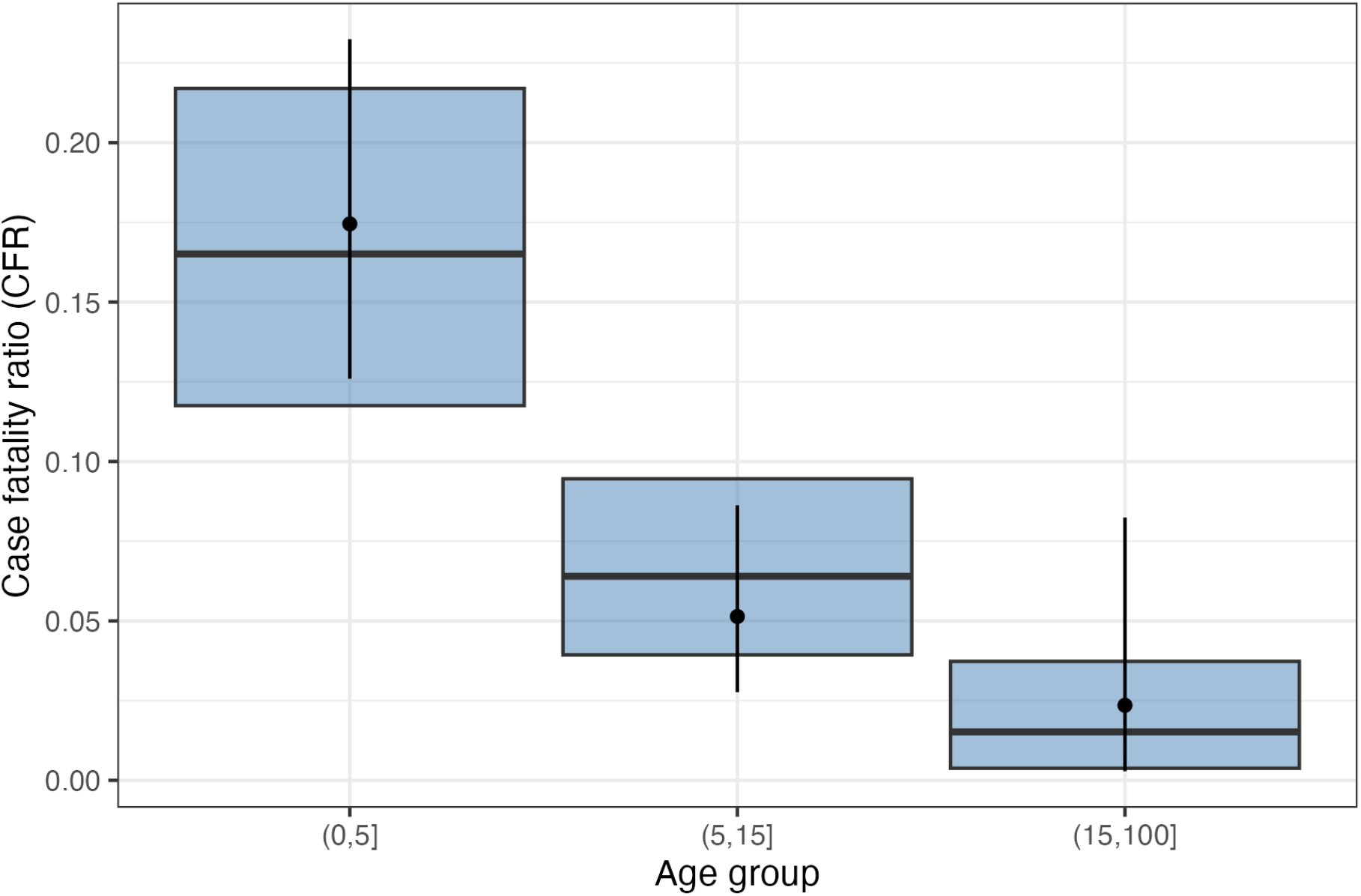
Observed vs expected case fatality ratio by age-group under best-fitting model to data extracted from the Bunge et al. systematic review, M7 (Hill function, age-only). Bars show mean and 95% credible intervals predicted by the model. Points and bars show the crude CFR calculated from the data and 95% binomial confidence intervals.

**Figure S2.**
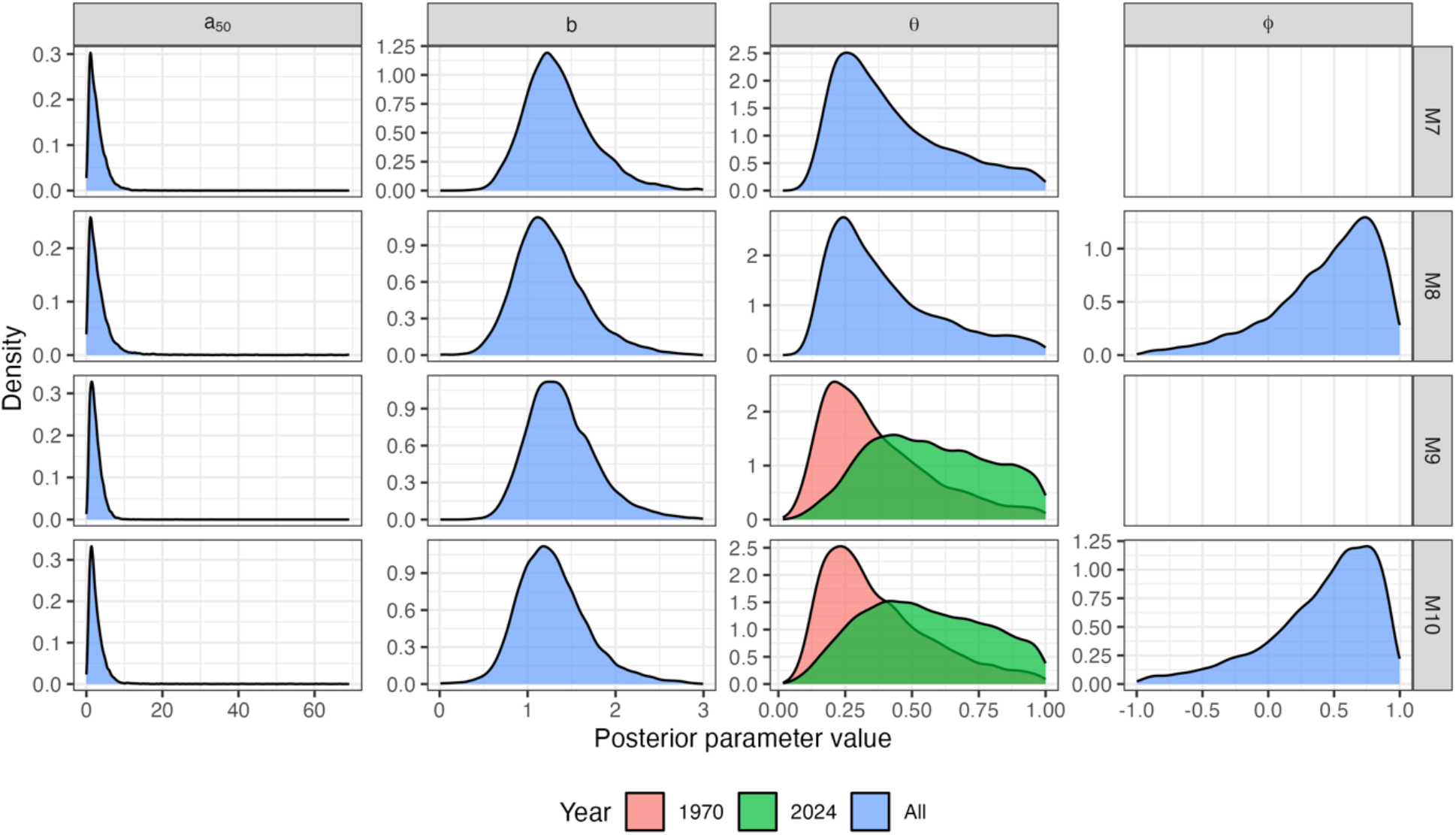
Posterior parameter densities for Hill function models of clade I CFR (M6-10) fitted to data extracted from the Bunge et al. systematic review.

**Table S1.** Description of fitted binomial regression models and posterior parameter estimates fitted to data reported in the Bunge et al systematic review (1), with difference in the expected log posterior density (ELPD) for each model compared to the best-fitting model (M7).

| M | Description \ Prior | $\theta_{1970}$ | $\theta_{2024}$ | $\phi$ | $a_{50}$ | $b$ | Difference<br>in ELPD |
| --- | --- | --- | --- | --- | --- | --- | --- |
| | | CFR at age zero<br>$\sim U(0, 1)$ | | Vaccine<br>effectiveness<br>against death<br>$\sim U(-1, 1)$ | Age of 50%<br>maximum CFR<br>$\sim U[0, 70]$ | Slope of Hill<br>function<br>$\sim U[0, 3]$ | Mean (s.e.) |
| 1 | Logistic, age only | 0.248<br>(0.098,0.414) |  |  | 7.133<br>(0.137,49.591) | 0.176<br>(0.053,0.746) | -8.9 (4.7) |
| 2 | No age, vaccination status only | 0.111<br>(0.085,0.142) |  | 0.808<br>(0.474,0.977) |  |  | -6.0 (6.5) |
| 3 | No age, year of onset only | 0.106<br>(0.066,0.151) | 0.081<br>(0.024,0.152) |  |  |  | -13.1 (7.8) |
| 4 | Logistic, age and vaccination status | 0.173<br>(0.089,0.360) |  | 0.703<br>(0.027,0.968) | 31.844<br>(0.431,68.388) | 0.777<br>(0.023,2.838) | -8.5 (5.7) |
| 5 | Logistic, age and year of onset | 0.247<br>(0.104,0.433) | 0.382<br>(0.103,0.819) |  | 3.696<br>(0.073,16.541) | 0.149<br>(0.059,0.252) | -8.9 (4.9) |
| 6 | Logistic, age, vaccination status, year of onset | 0.207<br>(0.086,0.394) | 0.258<br>(0.042,0.714) | 0.633<br>(-0.230,0.963) | 20.846<br>(0.127,67.362) | 0.526<br>(0.020,2.783) | -9.7 (5.7) |
| 7 | Hill, age only | 0.424<br>(0.154,0.928) |  |  | 2.860<br>(0.464,8.310) | 1.358<br>(0.721,2.275) | Ref |
| 8 | Hill, age and vaccination status | 0.400<br>(0.144,0.917) |  | 0.441<br>(-0.548,0.942) | 3.773<br>(0.424,14.037) | 1.261<br>(0.607,2.198) | -0.5 (1.4) |
| 9 | Hill, age and year of onset | 0.372<br>(0.107,0.892) | 0.570<br>(0.178,0.972) |  | 2.469<br>(0.519,6.326) | 1.390<br>(0.764,2.320) | -0.3 (0.9) |
| 10 | Hill, age, vaccination status, year of onset | 0.374<br>(0.112,0.881) | 0.548<br>(0.155,0.966) | 0.412<br>(-0.649,0.929) | 2.961<br>(0.459,8.334) | 1.290<br>(0.633,2.250) | -0.5 (1.6) |

### Sensitivity analysis: excluding ‘probable’ vaccinees

**Figure S3.**
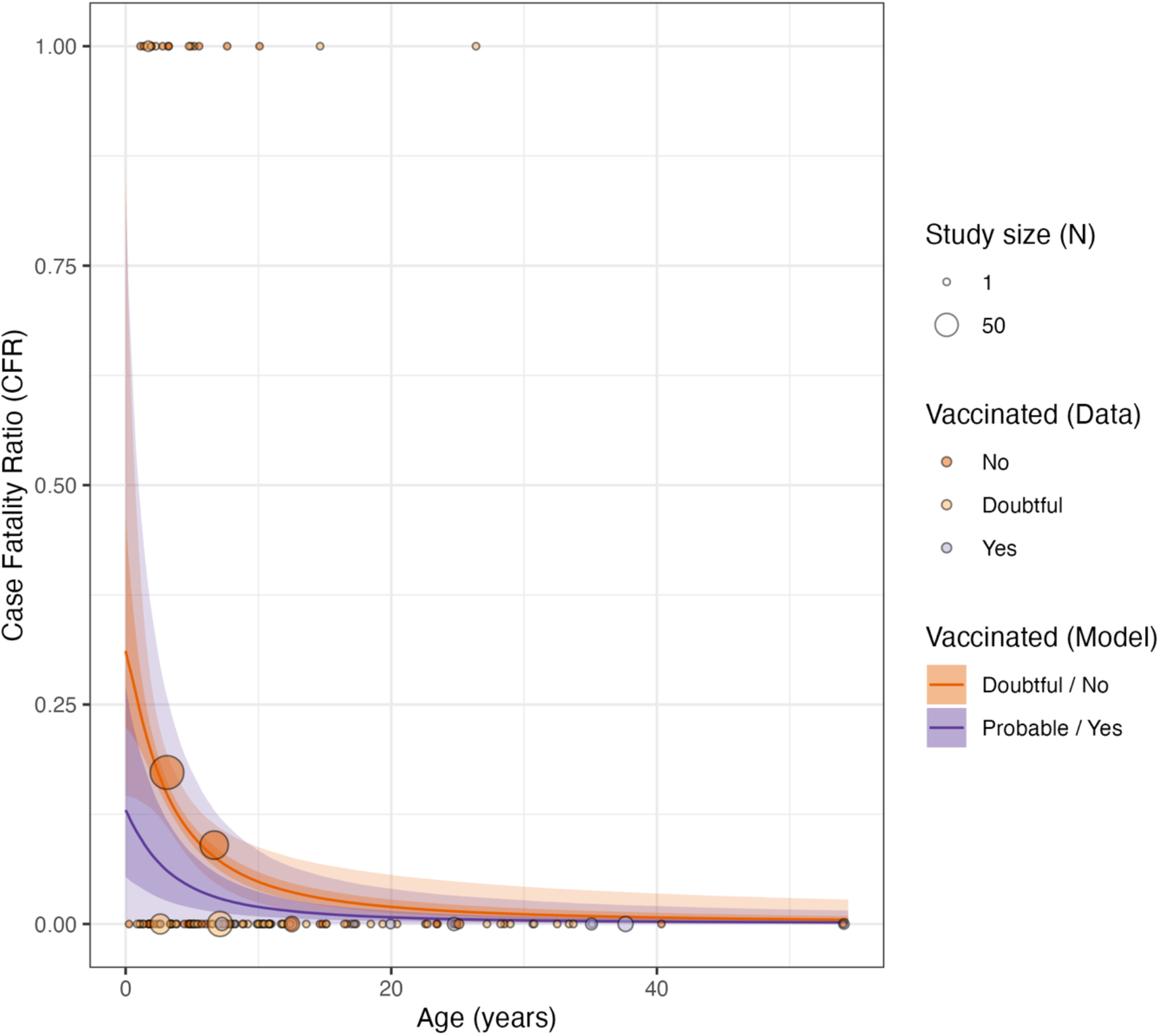
Estimated CFR by age and inferred vaccination status for mpox clade I under Model 8 (Hill, age and vaccination status) in the sensitivity analysis using data extracted from the Bunge et al. systematic review but excluding all cases born before 1980 whose vaccination status was unconfirmed (previously classified as ‘probable’ vaccinees). Solid lines depict the median posterior estimate for probable/vaccinated and doubtful/unvaccinated individuals, with shaded bands showing the 50% and 95% posterior predictive intervals. Points are coloured according to inferred vaccination status and show the outcome of individual cases where reported (0 = recovery, 1 = death). Larger points depict the crude CFRs calculated in studies reporting outcome by age-group (plotted according to the median age in the group, where reported and mid-point of the age range otherwise), with point size reflecting the number in each group.

**Figure S4.**
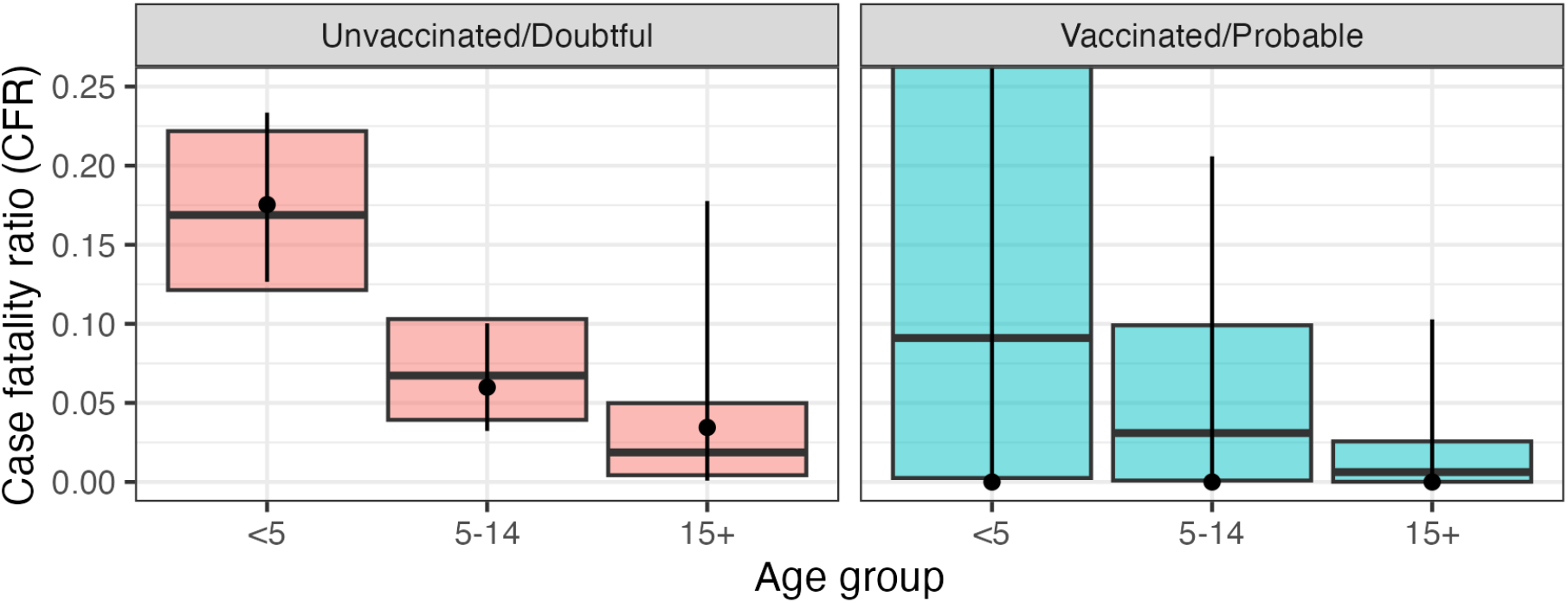
Observed vs expected case fatality ratio by age-group and vaccination status under best-fitting model (M8: Hill function, age and vaccination) in the sensitivity analysis using data extracted from the Bunge et al. systematic review but excluding all cases born before 1980 whose vaccination status was unconfirmed (previously classified as ‘probable’ vaccinees). Bars show mean and 95% credible intervals predicted by the model. Points and bars show the crude CFR calculated from the data and 95% binomial confidence intervals.

**Figure S5.**
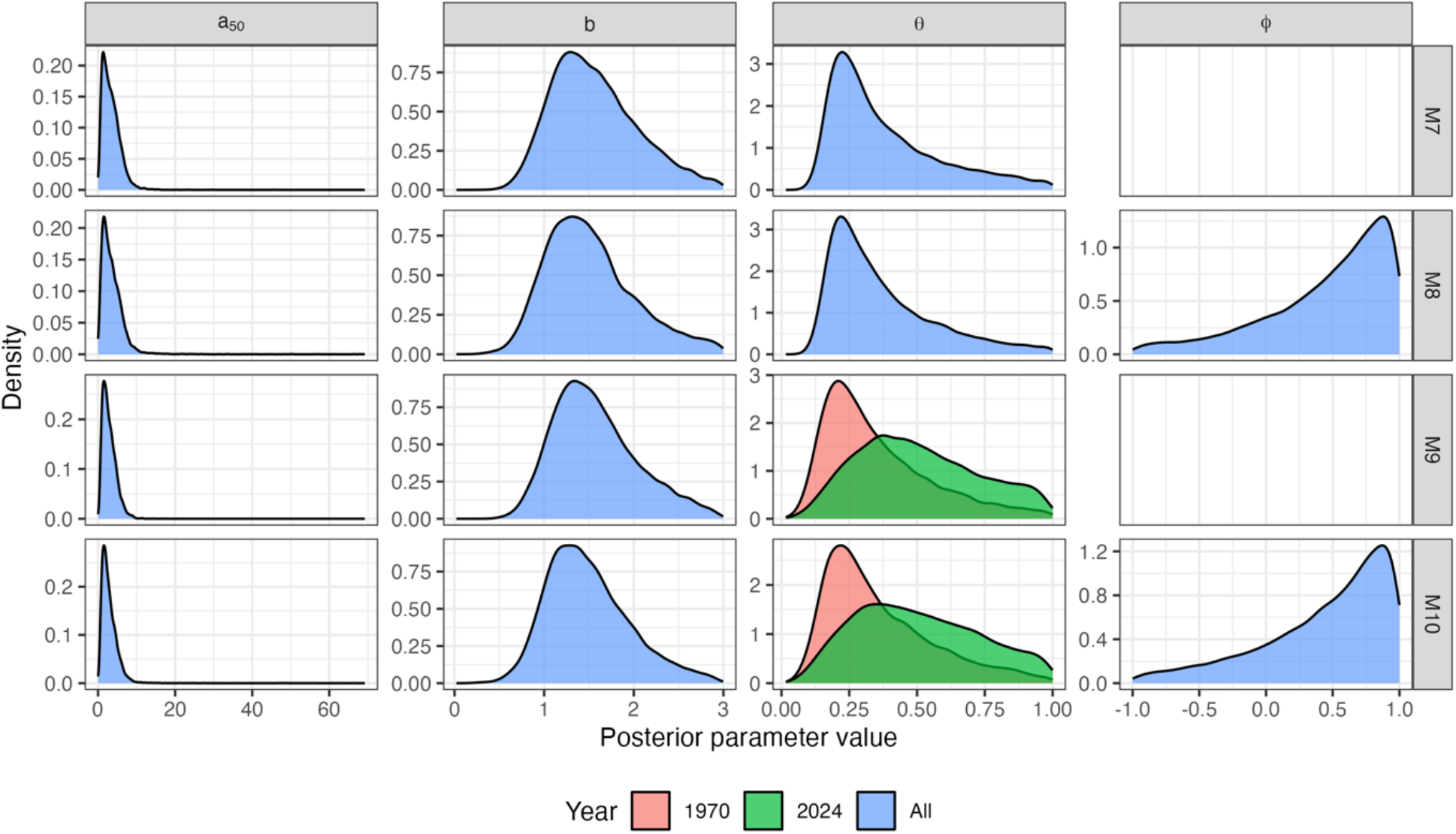
Posterior parameter densities for Hill function models of clade I CFR (M6-10) fitted to data extracted from the Bunge et al. systematic review but excluding all cases born before 1980 whose vaccination status was unconfirmed (previously classified as ‘probable’ vaccinees)

**Table S2.** Posterior parameter estimates for sensitivity analysis excluding DRC sitrep data and cases born before 1980 for whom vaccine status is unknown (assigned as ‘probable’ vaccinees in main analysis), with difference in the expected log posterior density (ELPD) for each model compared to the best-fitting model (M8).

| M | Description \ Prior | $\theta_{1970}$ | $\theta_{2024}$ | $\phi$ | $a_{50}$ | $b$ | Difference<br>in ELPD |
| --- | --- | --- | --- | --- | --- | --- | --- |
|  |  | CFR at age zero |  | Vaccine<br>efficacy<br>against death | Age of 50%<br>maximum CFR | Slope of Hill<br>function | Mean (s.e.) |
| | | $\sim U(0, 1)$ | | $\sim U(-1, 1)$ | $\sim U[0, 70]$ | $\sim U[0, 3]$ | |
| 1 | Logistic, age only | 0.326<br>(0.148,0.554) |  |  | 3.249<br>(0.102,12.104) | 0.225<br>(0.104,0.362) | -4.4 (3.2) |
| 2 | No age, vaccination status only | 0.111<br>(0.084,0.142) |  | 0.823<br>(0.335,0.995) |  |  | -7.4 (6.4) |
| 3 | No age, year of onset only | 0.117<br>(0.071,0.164) | 0.080<br>(0.020,0.161) |  |  |  | -12.9 (6.6) |
| 4 | Logistic, age and vaccination status | 0.301<br>(0.105,0.524) |  | 0.468<br>(-0.701,0.985) | 6.487<br>(0.091,48.082) | 0.307<br>(0.085,2.079) | -5.3 (3.3) |
| 5 | Logistic, age and year of onset | 0.304<br>(0.130,0.542) | 0.491<br>(0.139,0.938) |  | 2.316<br>(0.077,7.766) | 0.230<br>(0.114,0.353) | -4.4 (3.3) |
| 6 | Logistic, age, vaccination status, year of onset | 0.302<br>(0.123,0.538) | 0.464<br>(0.103,0.925) | 0.415<br>(-0.782,0.984) | 3.126<br>(0.066,13.365) | 0.235<br>(0.098,0.370) | -4.6 (3.3) |
| 7 | Hill, age only | 0.379<br>(0.150,0.890) |  |  | 3.400<br>(0.599,8.660) | 1.568<br>(0.808,2.660) | -0.4 (0.5) |
| 8 | Hill, age and vaccination status | 0.369<br>(0.147,0.879) |  | 0.454<br>(-0.747,0.984) | 3.715<br>(0.556,9.976) | 1.517<br>(0.741,2.714) | Ref |
| 9 | Hill, age and year of onset | 0.355<br>(0.106,0.869) | 0.508<br>(0.145,0.946) |  | 2.893<br>(0.629,6.999) | 1.574<br>(0.832,2.644) | -0.9 (0.8) |
| 10 | Hill, age, vaccination status, year of onset | 0.362<br>(0.110,0.859) | 0.505<br>(0.134,0.952) | 0.440<br>(-0.736,0.983) | 2.982<br>(0.600,7.612) | 1.493<br>(0.765,2.578) | -0.5 (0.7) |

### Analysis incorporating DRC sitrep data

**Figure S6.**
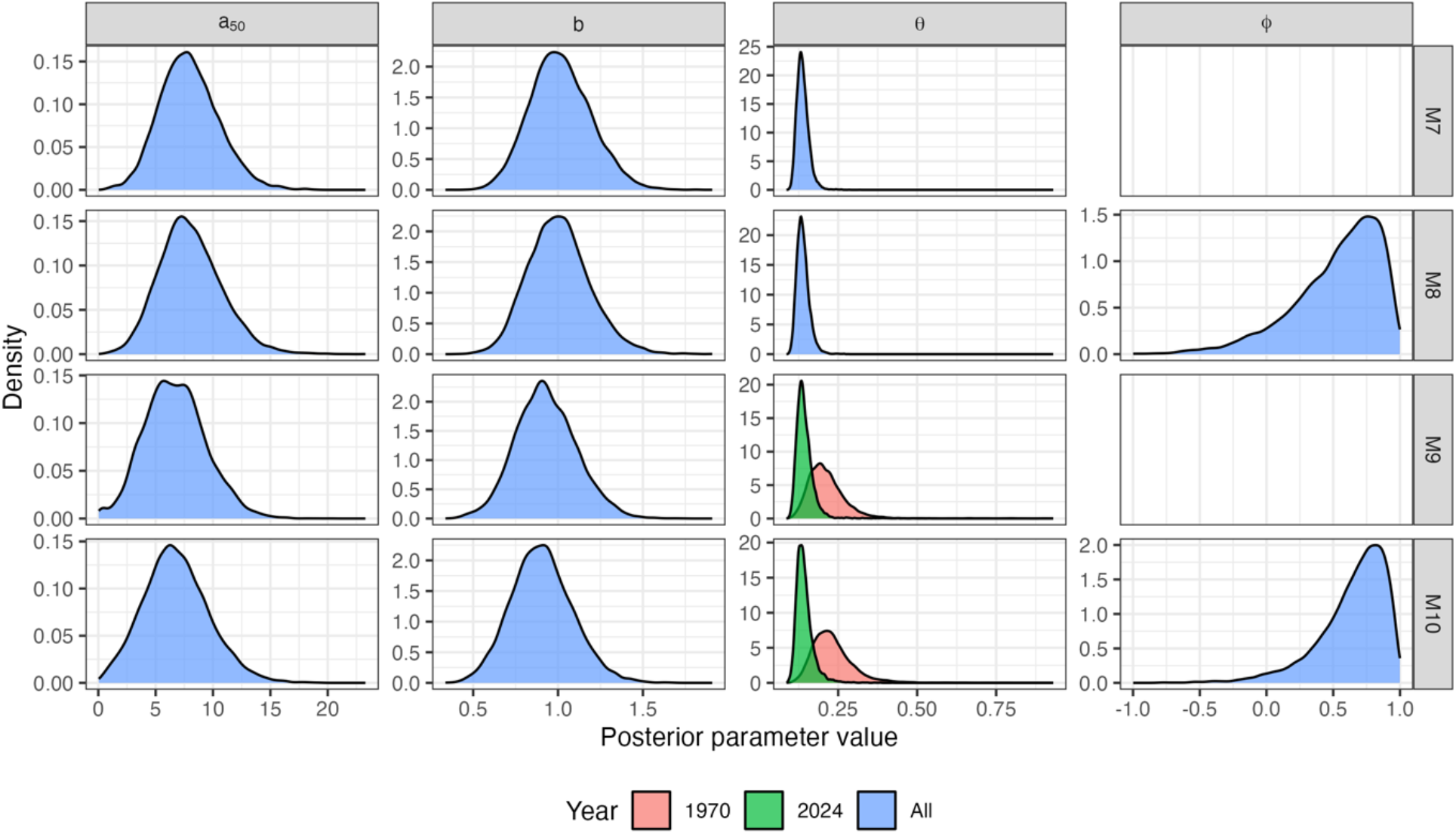
Posterior parameter densities for Hill function models of clade I CFR (M6-10) fitted to data data extracted from the Bunge et al. systematic review and additionally incorporating the DRC 2024 sitrep data.

**Figure S7.**
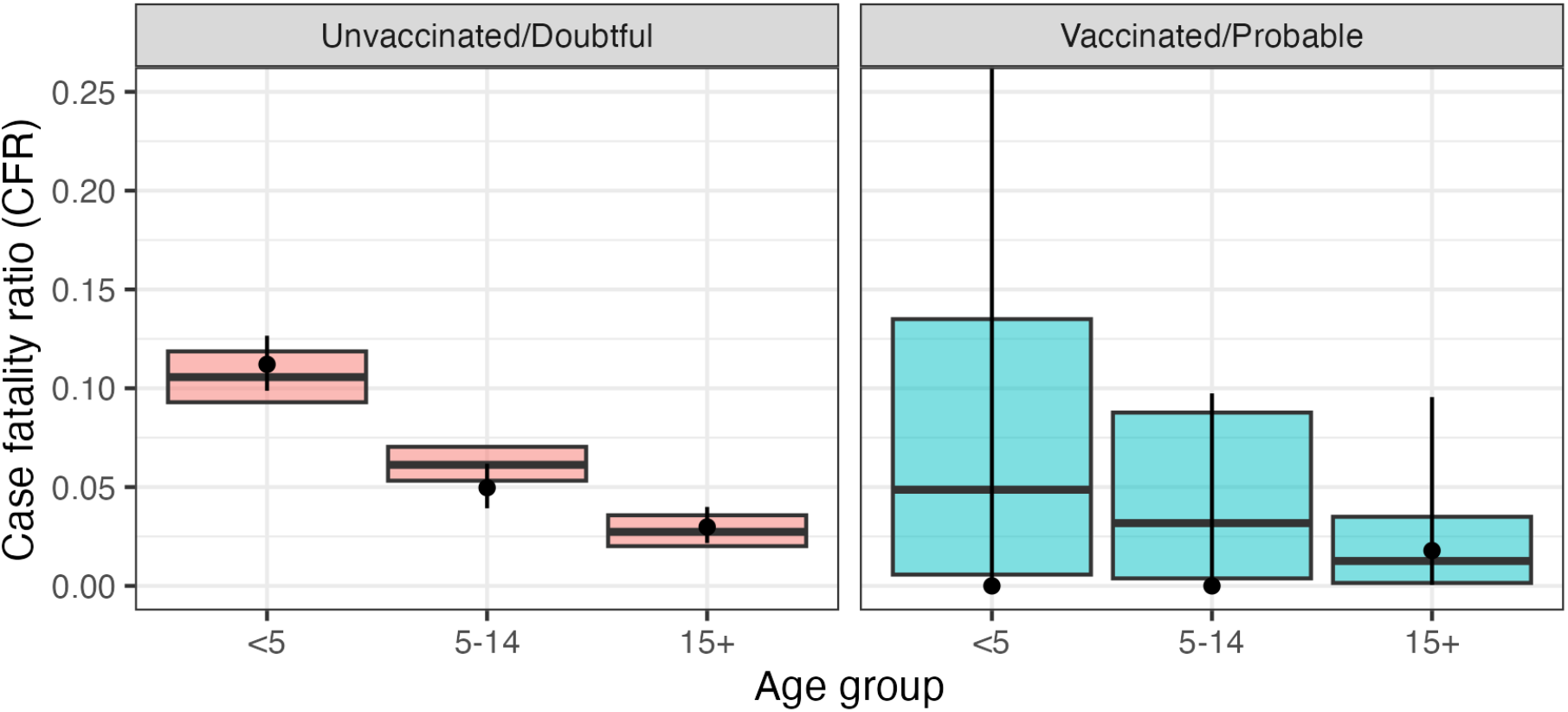
Observed vs expected case fatality ratio by age-group and vaccination status under best-fitting model (M10: Hill function, age, vaccination status, year of onset) based on data incorporating the DRC 2024 sitrep. Bars show mean and 95% credible intervals predicted by the model. Points and bars show the crude CFR calculated from the data and 95% binomial confidence intervals.

**Table S3.**
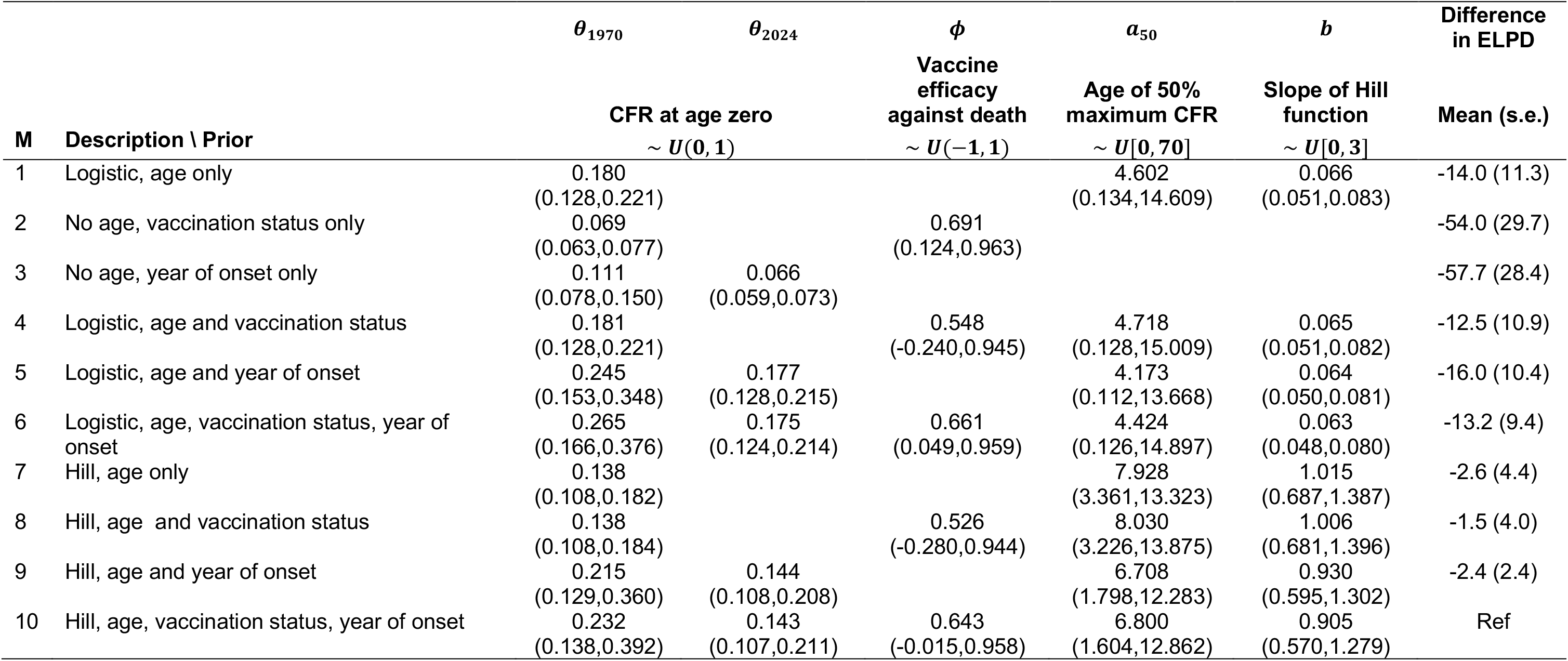
Description of fitted binomial regression models and posterior parameter estimates including DRC data, with difference in the 542 expected log posterior density (ELPD) for each model compared to the best-fitting model (M10).

### Sensitivity analysis: average age of cases in DRC sitrep data

**Figure S8.**
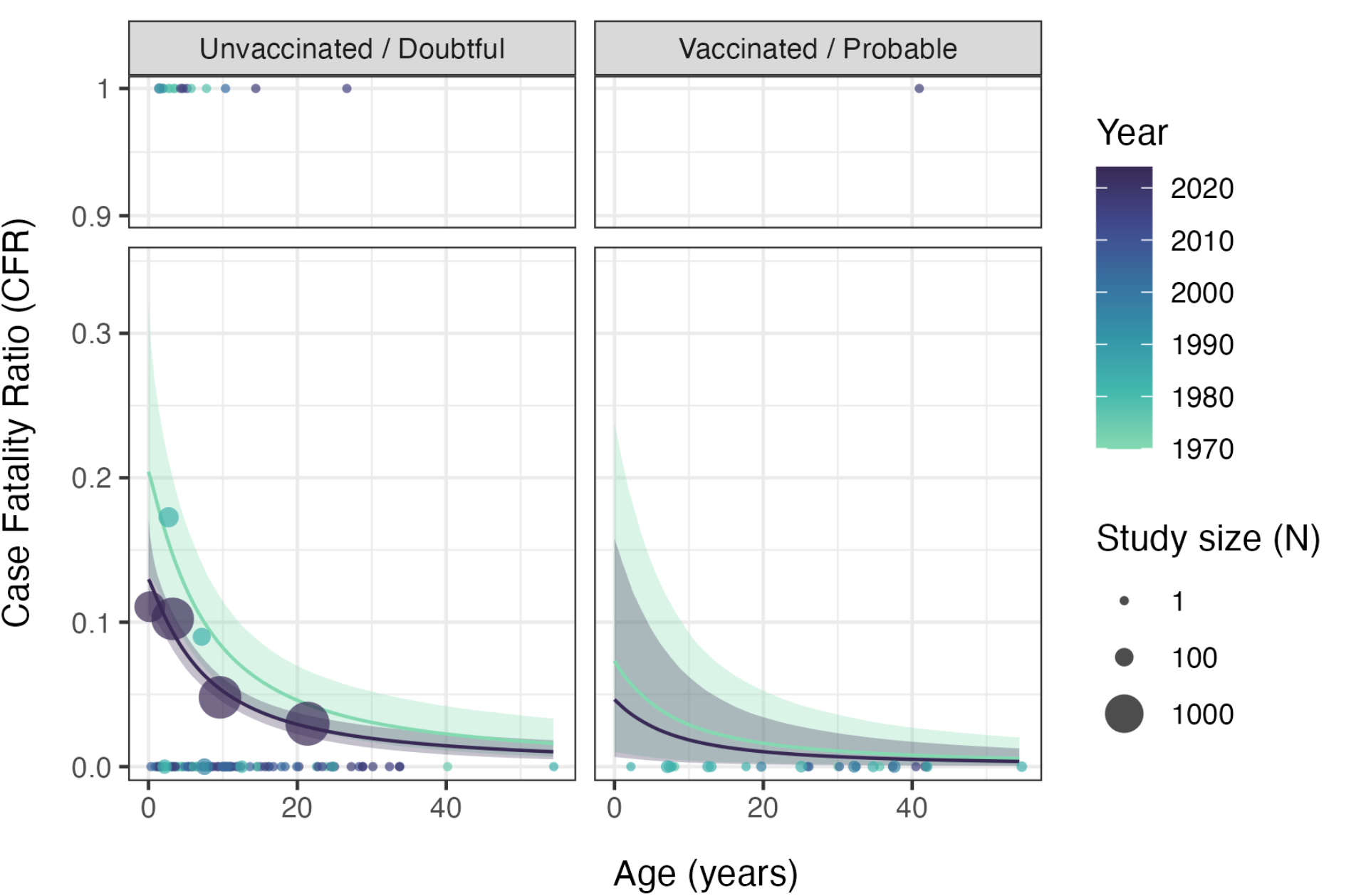
Estimated CFR by age, inferred vaccination status, and year of onset for mpox clade I under Model 10 (Hill, age, vaccination status, year) in the sensitivity analysis using data incorporating the 2024 DRC sitrep but assuming an average age of 21y among cases >15y. Solid lines depict the median posterior estimate for the posterior mean CFR at each age inferred for 1970 and 2024, with shaded bands showing the 50% and 95% credible intervals. Points are coloured according to year of outbreak and show the outcome of individual cases where reported (0 = recovery, 1 = death). Larger points depict the crude CFRs calculated in studies reporting outcome by age-group (plotted according to the median age in the group, where reported and mid-point of the age range otherwise), with point size reflecting the number in each group.

**Figure S9.**
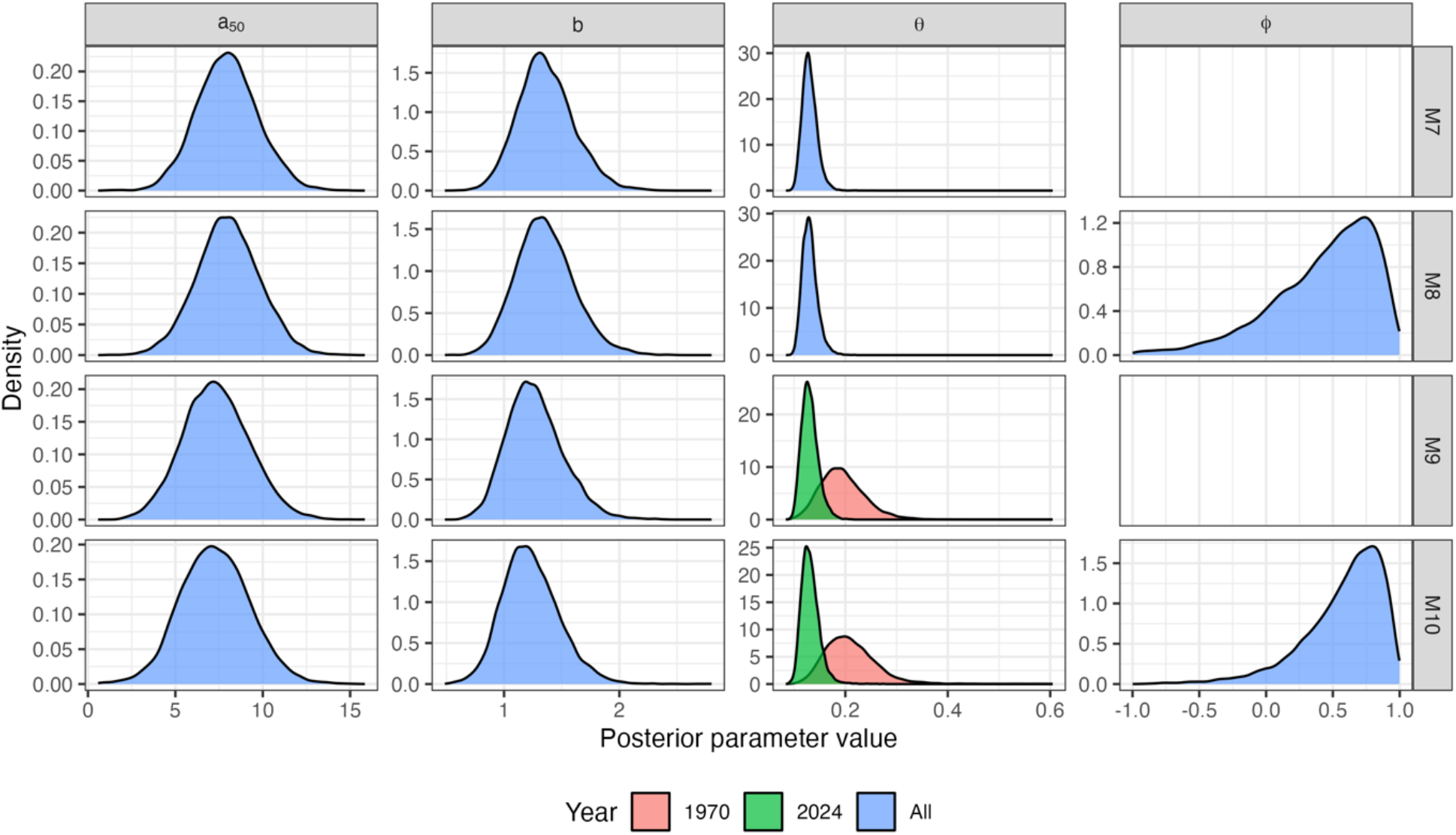
Posterior parameter densities for Hill function models of clade I CFR (M6-10) fitted to data data extracted from the Bunge et al. systematic review and additionally incorporating the DRC 2024 sitrep data, assuming an average age of 21y among cases >15y

**Figure S10.**
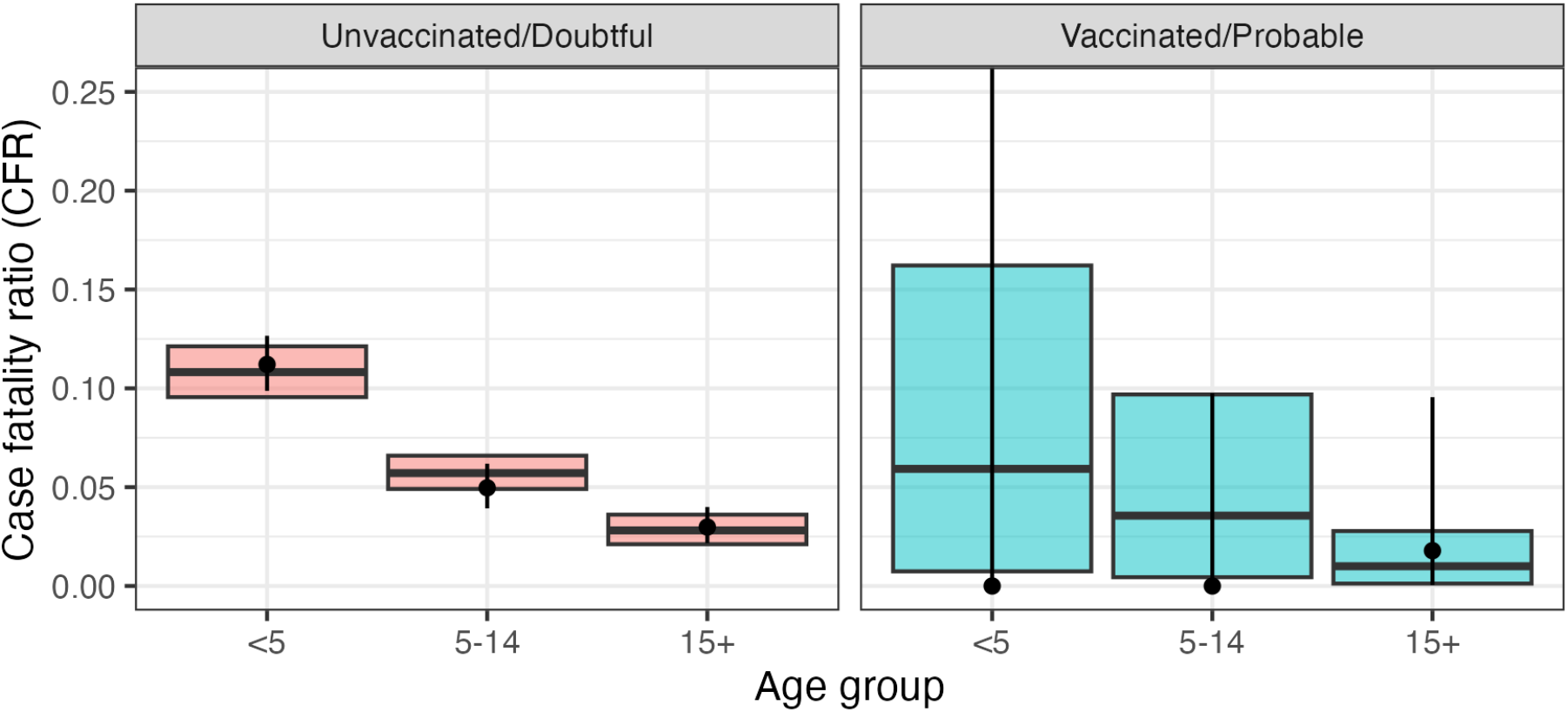
Observed vs expected case fatality ratio by age-group and vaccination status under best-fitting model (M10: Hill function, age, vaccination status and year) based on data incorporating the DRC 2024 sitrep, assuming an average age of 21y among cases >15y. Bars show mean and 95% credible intervals predicted by the model. Points and bars show the crude CFR calculated from the data and 95% binomial confidence intervals.

**Table S4.** Posterior parameter estimates including DRC sitrep data assuming an average age of 21y among cases >15y, with difference in the expected log posterior density (ELPD) for each model compared to the best-fitting model (M10).

| M | Description \ Prior | $\theta_{1970}$ | $\theta_{2024}$ | $\phi$ | $a_{50}$ | $b$ | Difference<br>in ELPD |
| --- | --- | --- | --- | --- | --- | --- | --- |
|  |  | CFR at age zero |  | Vaccine<br>efficacy<br>against death | Age of 50%<br>maximum CFR | Slope of Hill<br>function | Mean (s.e.) |
| | | $\sim U(0, 1)$ | | $\sim U(-1, 1)$ | $\sim U[0, 70]$ | $\sim U[0, 3]$ | |
| 1 | Logistic, age only | 0.202<br>(0.144,0.251) |  |  | 2.838<br>(0.086,8.626) | 0.106<br>(0.084,0.132) | -4.7 (6.4) |
| 2 | No age, vaccination status only | 0.069<br>(0.063,0.077) |  | 0.691<br>(0.124,0.963) |  |  | -57.7 (31.5) |
| 3 | No age, year of onset only | 0.111<br>(0.078,0.150) | 0.066<br>(0.059,0.073) |  |  |  | -61.4 (30.1) |
| 4 | Logistic, age and vaccination status | 0.202<br>(0.142,0.250) |  | 0.426<br>(-0.507,0.922) | 2.945<br>(0.089,9.112) | 0.106<br>(0.084,0.132) | -4.1 (6.3) |
| 5 | Logistic, age and year of onset | 0.281<br>(0.177,0.400) | 0.198<br>(0.143,0.243) |  | 2.588<br>(0.083,8.162) | 0.104<br>(0.083,0.128) | -5.3 (5.1) |
| 6 | Logistic, age, vaccination status, year of onset | 0.295<br>(0.183,0.421) | 0.196<br>(0.139,0.241) | 0.552<br>(-0.272,0.946) | 2.714<br>(0.089,8.553) | 0.103<br>(0.082,0.128) | -4.3 (5.0) |
| 7 | Hill, age only | 0.131<br>(0.106,0.163) |  |  | 7.970<br>(4.607,11.501) | 1.364<br>(0.928,1.881) | -0.9 (3.7) |
| 8 | Hill, age and vaccination status | 0.131<br>(0.107,0.165) |  | 0.428<br>(-0.516,0.931) | 8.012<br>(4.484,11.535) | 1.357<br>(0.911,1.893) | -0.2 (3.4) |
| 9 | Hill, age and year of onset | 0.195<br>(0.122,0.292) | 0.131<br>(0.104,0.169) |  | 7.348<br>(3.749,11.201) | 1.269<br>(0.853,1.800) | -0.4 (2.0) |
| 10 | Hill, age, vaccination status, year of onset | 0.210<br>(0.132,0.322) | 0.132<br>(0.105,0.172) | 0.579<br>(-0.186,0.947) | 7.274<br>(3.486,11.241) | 1.232<br>(0.800,1.766) | Ref |

## REFERENCES

1. Bunge EM, Hoet B, Chen L, Lienert F, Weidenthaler H, Baer LR, et al. The changing epidemiology of human monkeypox—A potential threat? A systematic review. PLoS Negl Trop Dis. 2022;16(2):1–20.

2. Deputy NP, Deckert J, Chard AN, Sandberg N, Moulia DL, Barkley E, et al. Vaccine Effectiveness of JYNNEOS against Mpox Disease in the United States. N Engl J Med. 2023 Jun 29;388(26):2434–43.

3. Taube JC, Rest EC, Lloyd-Smith JO, Bansal S. The global landscape of smallpox vaccination history and implications for current and future orthopoxvirus susceptibility: a modelling study. Lancet Infect Dis. 2022 Nov;S1473309922006648.

4. World Health Organization. 2022-23 Mpox (Monkeypox) Outbreak: Global Trends [Internet]. [cited 2023 Sep 14]. Available from: https://worldhealthorg.shinyapps.io/mpx_global/#2_Global_situation_update

5. Ulaeto D, Agafonov A, Burchfield J, Carter L, Happi C, Jakob R, et al. New nomenclature for mpox (monkeypox) and monkeypox virus clades. Lancet Infect Dis. 2023 Mar;23(3):273.

6. Mitjà O, Ogoina D, Titanji BK, Galvan C, Muyembe JJ, Marks M, et al. Monkeypox. Lancet Lond Engl. 2023 Jan 7;401(10370):60–74.

7. Mpox (monkeypox)-Democratic Republic of the Congo [Internet]. [cited 2023 Dec 5]. Available from: https://www.who.int/emergencies/disease-outbreak-news/item/2023-DON493

8. La variole simienne (monkeypox) en République démocratique du Congo: Rapport de la Situation Epidemiologique Sitrep No005 (du 18 - 24 mars 2024) - Democratic Republic of the Congo | ReliefWeb [Internet]. 2024 [cited 2024 Apr 10]. Available from: https://reliefweb.int/report/democratic-republic-congo/la-variole-simienne-monkeypox-en-republique-democratique-du-congo-rapport-de-la-situation-epidemiologique-sitrep-no005-du-18-24-mars-2024

9. Berthet N, Nakouné E, Whist E, Selekon B, Burguière AM, Manuguerra JC, et al. Maculopapular lesions in the Central African Republic. The Lancet. 2011 Oct 8;378(9799):1354.

10. Breman JG, Kalisa-Ruti, Steniowski MV, Zanotto E, Gromyko AI, Arita I. Human monkeypox, 1970-79. Bull World Health Organ. 1980;58(2):165–82.

11. Doshi RH, Guagliardo SAJ, Doty JB, Babeaux AD, Matheny A, Burgado J, et al. Epidemiologic and Ecologic Investigations of Monkeypox, Likouala Department, Republic of the Congo, 2017. Emerg Infect Dis. 2019 Feb;25(2):273–81.

12. Eltvedt AK, Christiansen M, Poulsen A. A Case Report of Monkeypox in a 4-Year-Old Boy from the DR Congo: Challenges of Diagnosis and Management. Case Rep Pediatr. 2020 Apr 9;2020:e8572596.

13. Jezek Z, Arita I, Mutombo M, Dunn C, Nakano JH, Szczeniowski M. Four generations of probable person-to-person transmission of human monkeypox. Am J Epidemiol. 1986 Jun;123(6):1004–12.

14. Formenty P, Muntasir MO, Damon I, Chowdhary V, Opoka ML, Monimart C, et al. Human Monkeypox Outbreak Caused by Novel Virus Belonging to Congo Basin Clade, Sudan, 2005. Emerg Infect Dis. 2010 Oct;16(10):1539–45.

15. Herve VMA, Belec L, Yayah G, Georges AJ. Monkeypox in central africa. About two strains isolated in Central African Republic. Med Mal Infect. 1989 May;19(5):322–4.

16. Kalthan E, Dondo-Fongbia JP, Yambele S, Dieu-Creer LR, Zepio R, Pamatika CM. Epidémie de 12 cas de maladie à virus monkeypox dans le district de Bangassou en République Centrafricaine en décembre 2015. Bull Société Pathol Exot. 2016 Dec 1;109(5):358–63.

17. Khodakevich L, Widy-Wirski R, Arita I, Marennikova SS, Nakano J, Meunier D. Monkey pox virus infection in humans in the Central African Republic. Bull Soc Pathol Exot Filiales. 1985 Jan 1;78(3):311–20.

18. Learned LA, Reynolds MG, Wassa DW, Li Y, Olson VA, Karem K, et al. Extended interhuman transmission of monkeypox in a hospital community in the Republic of the Congo, 2003. Am J Trop Med Hyg. 2005 Aug;73(2):428–34.

19. McCollum AM, Nakazawa Y, Ndongala GM, Pukuta E, Karhemere S, Lushima RS, et al. Human Monkeypox in the Kivus, a Conflict Region of the Democratic Republic of the Congo. Am J Trop Med Hyg. 2015 Oct 7;93(4):718–21.

20. Nakoune E, Lampaert E, Ndjapou SG, Janssens C, Zuniga I, Van Herp M, et al. A Nosocomial Outbreak of Human Monkeypox in the Central African Republic. Open Forum Infect Dis. 2017 Nov 3;4(4):ofx168.

21. Reynolds MG, Emerson GL, Pukuta E, Karhemere S, Muyembe JJ, Bikindou A, et al. Detection of Human Monkeypox in the Republic of the Congo Following Intensive Community Education. Am J Trop Med Hyg. 2013 May 1;88(5):982–5.

22. Jezek Z, Grab B, Szczeniowski M, Paluku KM, Mutombo M. Clinico-epidemiological features of monkeypox patients with an animal or human source of infection. Bull World Health Organ. 1988;66(4):459–64.

23. Mwanbal. Human Monkeypox -- Kasai Oriental, Zaire, 1996-1997 [Internet]. [cited 2023 Dec 5]. Available from: https://www.cdc.gov/mmwr/preview/mmwrhtml/00048673.htm

24. Jezek Z, Szczeniowski M, Paluku KM, Mutombo M. Human monkeypox: clinical features of 282 patients. J Infect Dis. 1987 Aug;156(2):293–8.

25. Bürkner PC. Bayesian Item Response Modeling in R with brms and Stan. J Stat Softw. 2021 Nov 30;100:1–54.

26. Carpenter B, Gelman A, Hoffman MD, Lee D, Goodrich B, Betancourt M, et al. Stan: A Probabilistic Programming Language. J Stat Softw. 2017 Jan 11;76:1–32.

27. Vehtari A, Gelman A, Gabry J. Practical Bayesian model evaluation using leave-one-out cross-validation and WAIC. Stat Comput. 2017;27(5):1413–32.

28. Paananen T, Piironen J, Bürkner PC, Vehtari A. Implicitly adaptive importance sampling. Stat Comput. 2021 Feb 9;31(2):16.

29. Ghani AC, Donnelly CA, Cox DR, Griffin JT, Fraser C, Lam TH, et al. Methods for estimating the case fatality ratio for a novel, emerging infectious disease. Am J Epidemiol. 2005;162(5):479–86.

30. Yinka-Ogunleye A, Dalhat M, Akinpelu A, Aruna O, Garba F, Ahmad A, et al. Mpox (monkeypox) risk and mortality associated with HIV infection: a national case-control study in Nigeria. BMJ Glob Health. 2023 Nov 30;8(11):e013126.

